# Haploinsufficiency of *ITSN1* is associated with Parkinson’s disease

**DOI:** 10.1101/2024.07.25.24310988

**Authors:** Thomas P. Spargo, Chloe F. Sands, Isabella R. Juan, Jonathan Mitchell, Vida Ravanmehr, Jessica C. Butts, Ruth B. De-Paula, Youngdoo Kim, Fengyuan Hu, Quanli Wang, Dimitrios Vitsios, Manik Garg, Mirko Messa, Guillermo del Angel, Daniel G. Calame, Hiba Saade, Laurie Robak, Ben Hollis, Huda Y. Zoghbi, Joshua Shulman, Slavé Petrovski, Ismael Al-Ramahi, Ioanna Tachmazidou, Ryan S. Dhindsa

**Affiliations:** Centre for Genomics Research, Discovery Sciences, R&D, AstraZeneca, Cambridge, UK; Department of Pathology and Immunology, Baylor College of Medicine, Houston, TX, USA; Department of Molecular and Human Genetics, Baylor College of Medicine, Houston, TX, USA; Department of Bioengineering, George R. Brown School of Engineering, Rice University, Houston, TX, USA; Quantitative and Computational Biology Program, Baylor College of Medicine, Houston, TX, USA; Centre for Genomics Research, Discovery Sciences, R&D, AstraZeneca, Waltham, MA, USA; Translational Genomics, Centre for Genomics Research, Discovery Sciences BioPharmaceuticals R&D, AstraZeneca, Gothenburg, Sweden; Centre for Genomics Research, Discovery Sciences, R&D, AstraZeneca, Boston, MA, USA; Department of Pediatrics, Texas Children’s Hospital, Houston, TX, USA; Department of Neurology, Baylor College of Medicine, Houston, TX, USA; Jan and Dan Duncan Neurological Research Institute, Texas Children’s Hospital, 1250 Moursund St., Suite N.1150, Houston, TX, 77030, USA; Howard Hughes Medical Institute, Baylor College of Medicine, Houston, TX, USA; Department of Neuroscience, Baylor College of Medicine, Houston, TX, USA; Center for Alzheimer’s and Neurodegenerative Diseases, Baylor College of Medicine, Houston, TX, USA; Department of Medicine, Austin Health, University of Melbourne, Melbourne, Australia

## Abstract

**Background:** Despite its significant heritability, the genetic underpinnings of Parkinson disease (PD) remain incompletely understood, particularly the role of rare variants. Advances in population-scale sequencing now provide an unprecedented opportunity to uncover additional large-effect rare genetic risk factors and expand our understanding of disease mechanisms.

**Methods:** We leveraged whole-genome sequence data with linked electronic health records from 490,560 UK Biobank participants, identifying 3,809 PD cases and 247,101 controls without a neurological disorder. We performed both variant-and gene-level association analyses to identify novel genetic associations with PD. We analyzed two additional independent case-control cohorts for replication (totaling 3,739 cases and 58,156 controls). Additionally, we performed functional validation of a novel PD association in a human synuclein-expressing *Drosophila* model.

**Findings:** In the UK Biobank, we replicated associations in well-established loci including *GBA1* and *LRRK2.* We also identified a novel association between protein-truncating variants (PTVs) in *ITSN1* and an increased risk of PD, with an effect size exceeding those of established loci (Fisher’s Exact Test: p=6.1x10^-7^; Odds ratio [95% confidence interval] = 10.53 [5.20, 21.34]). We replicated the *ITSN1* risk signal in a meta-analysis across all cohorts (Cochran-Mantel-Haenszel test p=5.7x10^-9^; Odds ratio [95% confidence interval] = 9.20 [4.66, 16.70]). In *Drosophila*, haploinsufficiency of the *ITSN1* ortholog (*Dap160*) exacerbated α-synuclein-induced compound eye degeneration and motor deficits.

**Interpretation:** We establish *ITSN1* as a novel risk gene for PD, with PTVs substantially increasing disease risk. *ITSN1* encodes a scaffold protein involved in synaptic vesicle endocytosis, a critical pathway increasingly recognized in PD pathogenesis. Our findings highlight the power of large-scale sequencing coupled with preclinical functional modeling to identify rare variant associations and elucidate disease mechanisms.

## Introduction

Parkinson’s disease (PD) is a heritable, age-related neurodegenerative disorder characterized by tremor at rest, rigidity, bradykinesia, loss of smell, and impaired gait/balance. Affecting 1-2% of adults aged over 65 years, PD is the second most common neurodegenerative disease.^1^ The neuropathological hallmarks of PD include the presence of intracellular inclusions containing aggregates of fibrillated α-Synuclein, known as Lewy bodies, and the degeneration of dopaminergic neurons in the substantia nigra pars compacta. Decreased dopamine release in the striatum and subsequent alterations in basal ganglia circuits are primarily responsible for PD motor manifestations.^2^ Current therapies provide symptomatic relief but are not curative, as they do not prevent dopaminergic neuron loss.

PD is thought to arise from a combination of environmental and genetic factors.^3^ Genetic discoveries in PD have uncovered key disease mechanisms, including mitochondrial and lysosomal dysfunction, impaired protein degradation,^4^ synaptic dysfunction, and disrupted vesicular trafficking. Initial discoveries emerged from investigating familial forms of the disease, which account for 5-10% of cases, and revealed rare highly penetrant alleles in several genes, such as *SNCA*, *PRKN*, and *LRRK2*. Attempts to identify genetic associations with sporadic PD have mostly focused on common variants through genome-wide association studies. These studies have revealed dozens of loci, some of which map to familial PD risk genes (e.g., *LRRK2* and *GBA1*). However, it has been challenging to glean biological insight from many of these common loci as they tend to be non-coding, confer small effect sizes, and can be confounded by linkage disequilibrium. Sequencing-based association studies allow the assessment of rare protein-coding variants with larger effect sizes and can offer clearer mechanistic and therapeutic insights. Although there have been recent strides in identifying rare variant loci in PD, the majority of PD heritability still remains unexplained.^3,5^

Here, we leveraged an unprecedented scale of genome sequencing (WGS) data with paired health record data from nearly 500,000 UK Biobank (UKB) participants to expand our understanding of the genetic architecture of PD. Through variant-and gene-level analyses, we replicated previously established risk loci and uncovered a novel loss-of-function signal in the endocytosis-related gene *ITSN1.* We replicated this link between *ITSN1* haploinsufficiency and PD in three additional case-control cohorts and performed functional validation in *Drosophila.* This study adds to the growing evidence that dysfunction of vesicle trafficking plays a crucial role in the pathophysiology of PD.

## Results

### Cohort characteristics and study design

We processed genome sequence data from 490,560 multi-ancestry UKB participants through our previously described cloud-based pipeline,^6^ removing samples with low sequencing quality, with low coverage depth, and index-sample pruning among closely related individuals (Methods). We then identified individuals with PD by aggregating self-reported data, hospital record-derived billing codes, and death registries (Methods). In total, we identified 3,809 PD cases in the UKB, including 3,702 of European ancestry, 69 of South Asian ancestry, 32 of African ancestry, and 6 of East Asian ancestry (**Fig. 1**).

**Figure 1.**
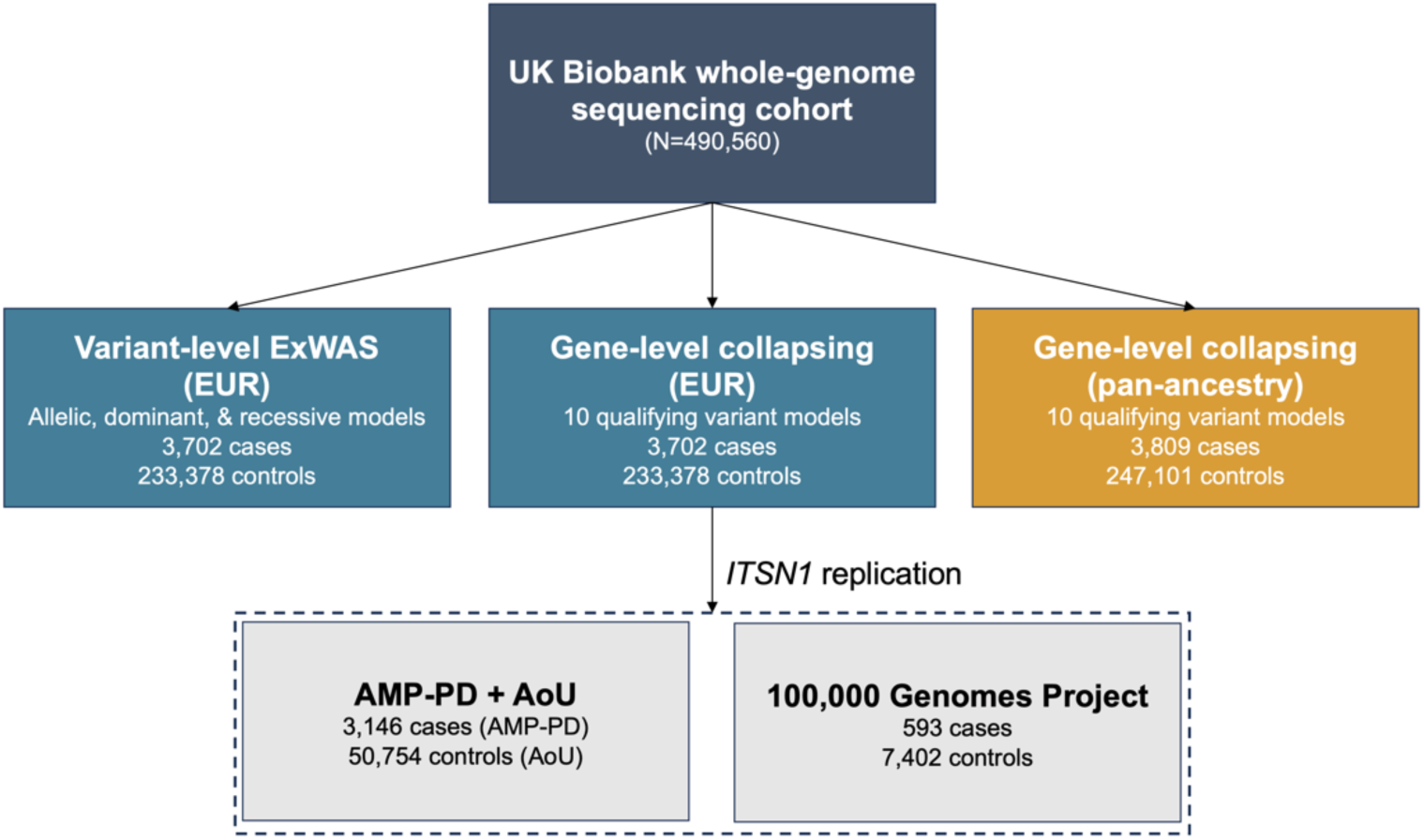
Genetic association study design. Schematic illustrating the genetic association analysis workflow. The discovery cohort included whole-genome sequenced participants from the UK Biobank. The pan-ancestry analysis included individuals of European (EUR), African, East Asian, and South Asian ancestries. The dotted box indicates the two additional EUR replication cohorts, which we used to replicate a novel association between *ITSN1* and PD. ExWAS = exome-wide association study; AMP-PD = Accelerating Medicines Partnership - Parkinson’s Disease; AoU = All of Us.

In European cases, the median age of PD diagnosis was 70.29 years (Interquartile range (IQR): 63.04-75.03), and 2,331 (63%) were male, consistent with epidemiological observations that PD is twice as prevalent in men.^7^ To reduce potential contamination from genetically related diagnoses, we restricted controls to participants without any diagnosis under ICD-10 Chapter VI (diseases of the nervous system).^6^ We then down sampled the remaining controls to match the 2:1 sex ratio observed in cases, resulting in 233,378 controls (**Table S1**).

To test for protein-coding associations with PD, we performed variant-level exome-wide association tests (ExWAS) and gene-level collapsing analyses (Methods). We carried out two versions of collapsing analysis: one restricted to individuals of non-Finnish European (NFE) ancestry (>90% of the UKB cohort) and one across all four ancestral groups in the UKB that had at least five PD cases (Methods). We performed gene-based replication analyses in two additional NFE cohorts. One comprised of 3,739 cases from the Accelerating Medicines Partnership program for Parkinson’s disease (AMP-PD) and 50,754 controls from All of Us (AoU). The other included 593 cases 7,402 controls from the 100,000 Genomes Project (100kGP) (**Fig. 1**; Methods). Associations from exome-wide discovery analyses were considered significant at p≤1x10^-8^ and suggestive at p≤1x10^-6^ (Methods).

### Variant-level associations

We first performed a variant-level ExWAS in the UKB EUR cohort to identify protein-coding variants associated with PD risk. Genomic inflation was well-controlled across the tested additive, dominant, and recessive genetic models (**Fig. S1**; λ_GC_ range: 1.01, 1.02). There were 36 significant (p*≤*1x10^-8^) variants across three independent loci (**Table 1**, **Table S2**). These results confirmed previously established associations, including the p.Gly2019Ser *LRRK2* variant (p=3.6x10^-12^; odds ratio (OR) [95% confidence interval (CI)] = 9.19 [5.67, 14.88]); the *MAPT* haplotype (p=3.2x10^-12^; OR [95% CI] = 0.79 [0.73, 0.84]); and the *MMRN1/SNCA* locus (p=4.0x10^-12^; OR [95% CI] = 1.41 [1.28, 1.55]), (**Fig. 2A**, **Fig. S2**).^8^ We also detected five established pathogenic *GBA1* variants that fell short of the study-wide significance threshold, including the p.Glu365Lys association (1-155236376-C-T, p= 4.47x10^-8^, OR [95% CI] = 1.61 [1.37, 1.89]) and others (**Table S2**).^9^

**Figure 2.**
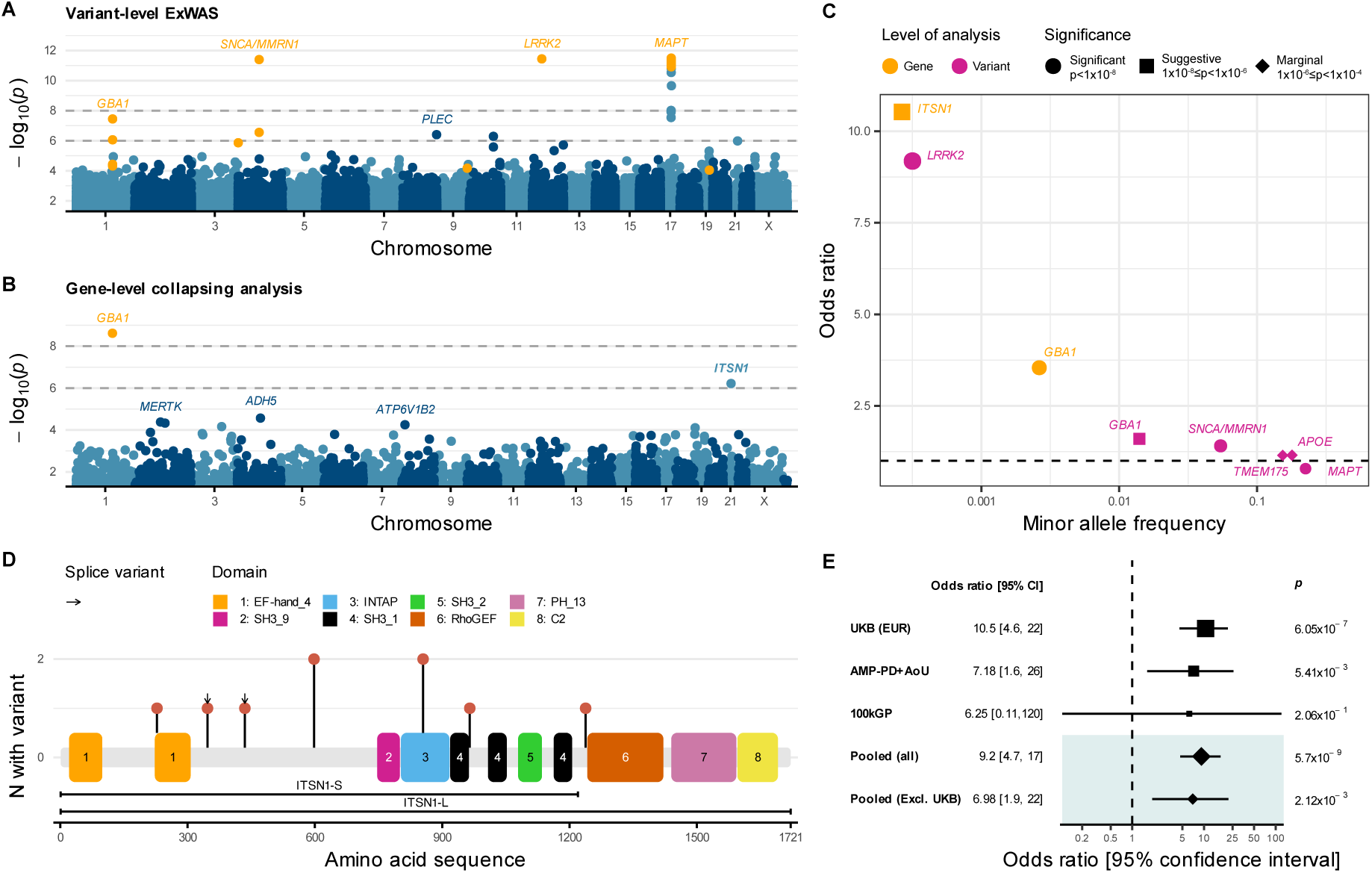
Variant-and gene-level associations with PD. (**A, B**) Manhattan plots of variant-level (A) and gene-level (B) associations with PD in UKB Europeans. For each plot, if the same association was detected in multiple models, we depict the most significant association. See Figures S2, S4 for per-model plots. Horizontal dashed lines indicate the significance (p<1x10^-8^) and suggestive (p<1x10^-6^) thresholds. Orange points indicate marginally significant (p<1x10^-4^) associations in genes previously implicated in PD. (**C**) Odds ratio versus minor allele frequency for *ITSN1* and other established PD loci that achieved p<1x10^-4^ in the ExWAS or collapsing analysis. (**D**) Lollipop plot showing the location of *ITSN1* PTVs in cases against the MANE transcript (ENST00000381318) and PFam domains of the corresponding protein (Q15811); bars below the transcript indicate the ITSN1-L and ITSN1-S isoforms. (**E**) Analyses of the association between *ITSN1* and PD under the ptv collapsing-analysis model across the UKB discovery and replication cohorts, ‘pooled’ association statistics, shaded gray, are based on cohort-stratified exact Cochran-Mantel-Haenszel tests. UKB = UK Biobank; EUR = European-ancestry cohort; AMP-PD = Accelerating Medicines Partnership - Parkinson’s Disease; AoU = All of Us; 100kGP = 100,000 Genomes Project.

**Table 1.**
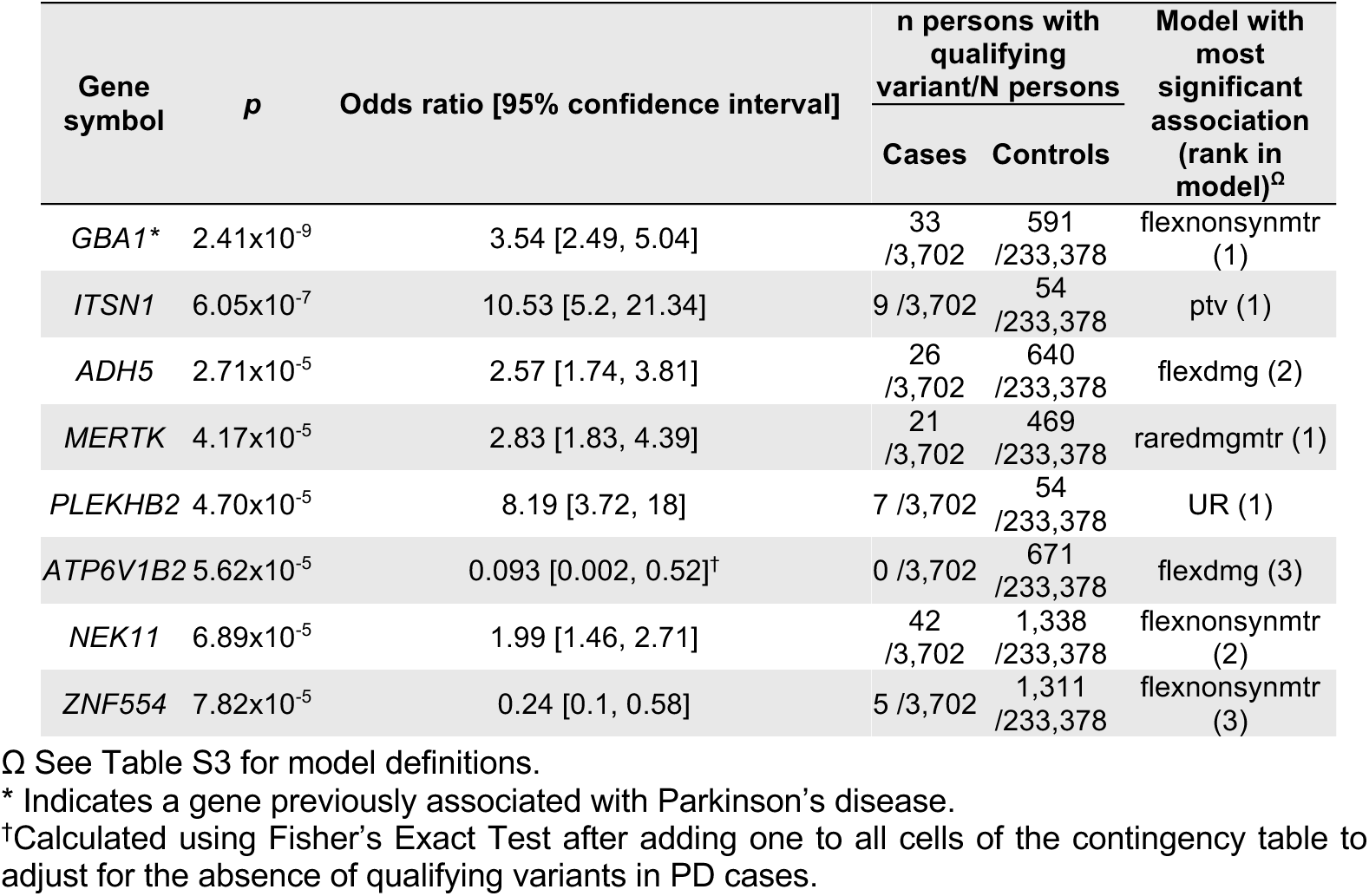
Top gene-level associations with Parkinson’s disease. Gene-level associations that achieved p<1x10^-4^ across the nine non-synonymous collapsing models. The study-wide significance threshold is p<1x10^-8^.

| Gene symbol | $p$ | Odds ratio [95% confidence interval] | n persons with qualifying variant/N persons | | Model with most significant association (rank in model)<sup>Ω</sup> |
| --- | --- | --- | --- | --- | --- |
|  |  |  | Cases | Controls |  |
| *GBA1* * | $2.41 \times 10^{-9}$ | 3.54 [2.49, 5.04] | 33 / 3,702 | 591 / 233,378 | flexnonsynmtr (1) |
| *ITSN1* | $6.05 \times 10^{-7}$ | 10.53 [5.2, 21.34] | 9 / 3,702 | 54 / 233,378 | ptv (1) |
| *ADH5* | $2.71 \times 10^{-5}$ | 2.57 [1.74, 3.81] | 26 / 3,702 | 640 / 233,378 | flexdmg (2) |
| *MERTK* | $4.17 \times 10^{-5}$ | 2.83 [1.83, 4.39] | 21 / 3,702 | 469 / 233,378 | raredmgmtr (1) |
| *PLEKHB2* | $4.70 \times 10^{-5}$ | 8.19 [3.72, 18] | 7 / 3,702 | 54 / 233,378 | UR (1) |
| *ATP6V1B2* | $5.62 \times 10^{-5}$ | 0.093 [0.002, 0.52]<sup>†</sup> | 0 / 3,702 | 671 / 233,378 | flexdmg (3) |
| *NEK11* | $6.89 \times 10^{-5}$ | 1.99 [1.46, 2.71] | 42 / 3,702 | 1,338 / 233,378 | flexnonsynmtr (2) |
| *ZNF554* | $7.82 \times 10^{-5}$ | 0.24 [0.1, 0.58] | 5 / 3,702 | 1,311 / 233,378 | flexnonsynmtr (3) |

<sup>Ω</sup> See Table S3 for model definitions.
\* Indicates a gene previously associated with Parkinson's disease.
<sup>†</sup> Calculated using Fisher's Exact Test after adding one to all cells of the contingency table to adjust for the absence of qualifying variants in PD cases.

### Gene-level collapsing analysis

We have previously introduced a gene-level collapsing analysis framework that tests rare variants in aggregate, which demonstrably bolsters power for rare variant-driven associations.^6,10,11^ In this method, we identify rare variants that meet a predefined set of criteria (that is, ‘qualifying variants’ or ‘QVs’) in each gene and test for their aggregate effect. We tested a total of 18,930 genes under nine different non-synonymous QV models to evaluate a range of possible genetic architectures (**Table S3**). There was no evidence of genomic inflation (**Fig. S3**; median λ_GC_ = 1.01; range: 0.98, 1.03).

*GBA1* achieved study-wide significance in the UKB European analysis (**Table 1**, **Fig. 2B**, **Fig. S4, Table S4**). This association was most significant under the *flexnonsynmtr* QV model, which considers rare (minor allele frequency (MAF) <0.1%) protein-truncating variants and missense variants that fall in missense-intolerant regions of the gene (p=2.4x10^-9^, OR [95% CI]: 3.54 [2.49, 5.04]). The fact that *GBA1* reached significance in the gene-level collapsing tests but not the ExWAS demonstrates the power gained by aggregating the contributions of multiple distinct rare variants within a gene. We performed a “leave-one-out” to identify which individual variants contributed most to the association under the *flexnonsynmtr* model. This analysis identified the variants 1-155236366-C-T (p.Arg368His), 1-155238260-G-C (p.Ser212*), and 1-155240629-C-T (c.115+1G>A) as most influential (**Fig. S5**; **Table S5**). Notably, only two out of five ExWAS associations (p<1x10^-4^) were included as QVs in the *flexnonsynmtr* model; the three excluded variants, including two previously implicated in PD (1-155236376-C-T [p.Glu365Lys] and 1-155236246-G-A [p.Thr408Met]), had MAFs exceeding the *flexnonsynmtr* model threshold (**Table S2**).

The second ranking gene in the collapsing analysis was *ITSN1*, which had a suggestive (p=6.1x10^-7^) association under the protein-truncating variant (*ptv*) model. *ITSN1* PTVs conferred a striking effect size: carriers had 10 times greater odds of developing PD than non-carriers (OR [95% CI] = 10.53 [5.2, 21.34]). *ITSN1* PTVs were rare, with a collective case carrier frequency of 0.24% (n=9/3,702) compared to a control carrier frequency of 0.02% (n=54/233,378). Though the confidence intervals overlap, the effect size for the *ITSN1* association exceeds those of previously established loci (**Fig. 2C**), including the deleterious *LRRK2* p.Gly2019Ser variant.

Several other top-ranking hits in the European collapsing analysis that did not meet our stringent p-value cutoff had prior functional or genetic support. These include *ADH5*, which encodes alcohol dehydrogenase, and *MERTK*, which has been implicated in the mediation of alpha-synuclein fibril update by human microglia (**Table 1**, **Table S4**).^12^ However, additional analyses are required to firmly implicate these genes as PD risk genes.

Finally, we performed a pan-ancestry analysis collapsing analysis, which recapitulated the top-ranking hits from the European ancestry group analysis (**Table S6)**. No additional genes reached significance. However, future studies incorporating more cases from diverse ancestries will be crucial for further uncovering the genetic architecture of PD and ensuring equitable advances in genetic diagnoses and targeted therapies.

### *In silico* investigation of *ITSN1* variation

Qualifying *ITSN1* PTVs were observed in 0.24% of cases compared to 0.023% of controls. Among UKB Europeans prior to downsampling of female controls, PD occurred with similar frequency in males (5/44; 11.3%) and females (4/36; 11.1%) with qualifying *ITSN1* PTVs. In total, there were seven unique qualifying PTVs among UKB European cases; five were singletons and two were observed in two cases each (21-33782101-C-T [p.Gln598*] and 21-33811223-G-T [c.2567+1G>T]) (**Fig. 2D**; **Table S7**). These PTVs were scattered throughout the *ITSN1* gene body, suggestive of a loss-of-function mechanism (**Fig. 2D**). There are 16 impacted protein-coding *ITSN1* transcripts in Ensembl (v92). Given the level of isoform diversity, we ran collapsing analysis on each transcript individually. There was no preferential enrichment of the collapsing signal amongst any particular transcript. Crucially, the signal persisted in transcripts most highly expressed in the nervous system (**Table S10; Fig. S7**).

Consistent with the UK Biobank control frequency, the cumulative PTV frequency is 0.021% among non-Finnish European non-neurological controls in gnomAD v2.1.1 (n=44,779 exomes) (**Table S8, S9**). We next queried our prior phenome-wide association study (PheWAS; azphewas.com)^6^ to determine if PTVs in this gene were associated with any other phenotype in the UK Biobank. Among the ∼18,000 tested phenotypes, none were associated with *ITSN1* at p<1x10^-4^. It is notable, however, that haploinsufficiency of *ITSN1* has recently been associated with autism spectrum disorder (ASD).^13^

### Replication analysis

We next aimed to replicate the *ITSN1* signal in two independent case-control datasets. Given the rarity of *ITSN1* PTVs, we constructed a maximally powered primary replication dataset (see Methods) by combining 3,146 AMP-PD cases with 50,754 non-neurological controls in All of Us (AoU). Under the *ptv* model, *ITSN1* was significantly associated with increased PD risk (p=5.4x10^-3^; OR [95% CI] = 7.18 [1.61, 25.75]). QVs were present in 4/3,146 (0.13%) AMP-PD cases and 9/50,754 (0.02%) AoU controls (**Table S7**). The cumulative frequency of PTVs was consistent between the controls from UK Biobank (0.02%) and AoU (0.02%). The effect size was in the same direction in the considerably smaller 100kGP replication cohort, which was underpowered to detect a significant effect in isolation (**Fig. 2E**, see Supp. Methods and **Fig. S8** for power analysis). An exact Cochran-Mantel-Haenszel (CMH) test meta-analysis across these two replication cohorts achieved a p=2.1x10^-3^ (OR [95% CI] = 6.98 [1.89, 21.85]). Finally, taking the combined evidence from the UKB and these independent replication cohorts, the *ITSN1* signal became statistically unequivocal (CMH p=5.7x10^-9^; OR [95% CI] = 9.20 [4.66, 16.70]). These analyses firmly link *ITSN1* haploinsufficiency with a substantial increased risk of PD.

### Age-of-diagnosis

Strongly genetic forms of disease can often present with earlier age of onset and resistance to treatment.^1,3^ We thus compared the age at diagnosis for *LRRK2, GBA,* and *ITSN1* variant carriers to the remaining PD cases in the UK Biobank European ancestry cohort. The remaining cohort had a median age of diagnosis of 70.3 (IQR: 63.1, 75.1). In contrast, the median age of diagnosis was 66.1 years (IQR: 59.7, 71.5) in the 33 GBA QV carriers (*flexnonsynmtr* collapsing model) and 66 (IQR: 61.3, 71.4) in the 19 p.Gly2019Ser *LRRK2* variant carriers (p_GBA_=0.013; p_LRRK2_=0.061; **Fig. S6**). The age of diagnosis also trended earlier for the nine *ITSN1* PTV carriers (median = 63.5, IQR: 59.0, 73.7; p=0.054; **Fig. S6**). These data suggest that PTVs in *ITSN1* may predispose individuals to an earlier onset of disease, but further deep clinical characterization of additional *ITSN1* PTV carriers is required to validate these findings and better understand potential variations in disease progression.

### Supporting evidence for *ITSN1* as a PD risk gene

We next analyzed protein-protein interaction (PPI) networks in STRING-db,^14^ which revealed that ITSN1 physically interacts with several critical endocytic proteins (**Fig 3A, B**). This is consistent with ITSN1’s established role as a multivalent scaffold protein that organizes endocytic protein interaction networks to promote clathrin-mediated endocytosis and synaptic vesicle recycling.^15^ Interestingly, four of ITSN1’s direct interactors have been linked to PD: Synaptojanin-1 (*SYNJ1*) and Auxilin (*DNAJC6*) are firmly associated with early-onset PD whilst Endophilin-A1 (*SH3GL2*) and Actin nucleation-promoting factor WASL (*WASL*) have recently emerged as candidate PD genes.^16^ Moreover, ITSN1’s second-degree neighbors were significantly enriched for other PD genes (n=11 genes, Fisher’s exact test p<1x10^-5^; **Fig. 3B**). Many of these proteins are involved in the downstream endolysosomal pathway. Overall, ITSN1 appears to be embedded in a dense, interconnected network of PD-related proteins that cooperate at virtually every stage of vesicular transport (**Fig. 3A**).

**Figure 3.**
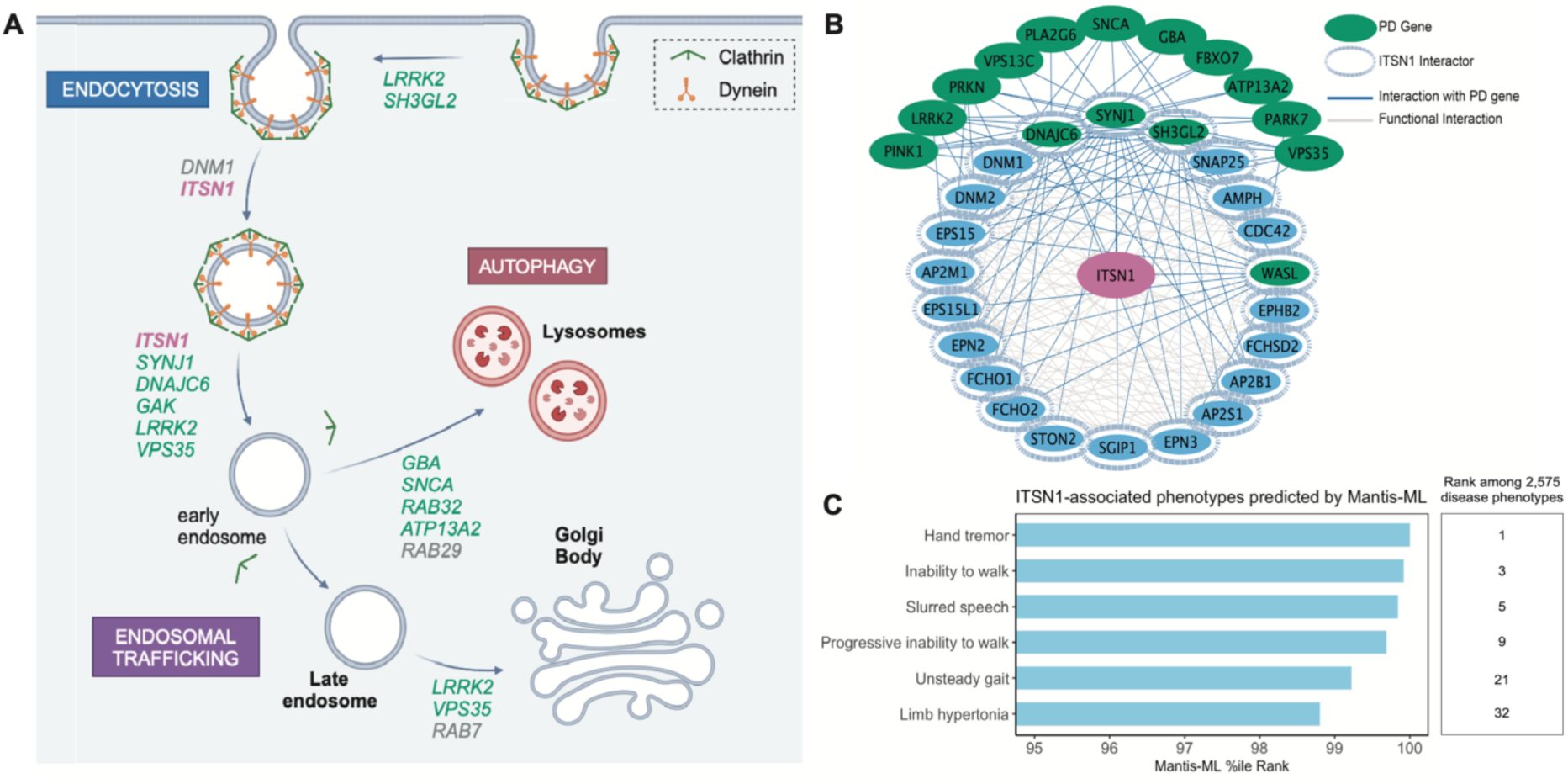
Interrogation of ITSN1’s function and connection to PD. (**A**) Schematic of vesicular trafficking pathway, with ITSN1 listed in red and known PD genes in green. (**B**) Protein-protein interaction map of ITSN1 (center) and 20 of its top interactors (inner ring), as well as previously identified PD genes.^18,19^ (**C**) Disease phenotypes predicted to be associated with ITSN1 by the machine learning algorithm Mantis-ML. All shown are well-established clinical signs of parkinsonism and were ranked by Mantis-ML in the top 1.25% of predicted associations.

We next tested whether there was orthogonal evidence to support *ITSN1’*s association with PD using Mantis-ML.^17^ This machine-learning framework predicts genotype-phenotype relationships using a vast set of features, including gene expression data, genetic intolerance, PPI networks, and a knowledge graph with over 8.7M edges. Remarkably, across 2,575 studied phenotypes, “hand tremor”, “inability to walk”, and “slurred speech” were among the top 10 Mantis-ML predictions for *ITSN1-*related phenotypes (**Fig. 3C** and **Table S11**). Collectively, these *in silico* analyses further support the link between *ITSN1* and PD-related biology.

### *In vivo* investigation of *ITSN1*

We and others have used transgenic *Drosophila* lines expressing human α-Synuclein to study how genes associated with PD interact with α-synuclein-mediated neurotoxic mechanisms. Here, we used a well-validated PD model^20,21^ to functionally characterize the effects of *ITSN1* haploinsufficiency and investigate its potential interaction with α-Synuclein. *Drosophila* expressing α-Synuclein exhibit photoreceptor degeneration, which appears as disorganization of the external hexagonal facets (**Fig. 4A, B**). This "rough eye" phenotype has been observed in *Drosophila* models of neurodegeneration, including Alzheimer’s disease and PD.^22,23^ Flies carrying a heterozygous null variant in *Dap160* (the *Drosophila* ortholog of *ITSN1*)^24^ had exacerbated α-Synuclein-induced degeneration, with more severe disorganization covering a larger eye area (**Fig. 4C, E**). Strikingly, overexpression of *Dap160* ameliorated the α-Synuclein-induced rough eye phenotype (**Fig. 4D, E**).

**Figure 4.**
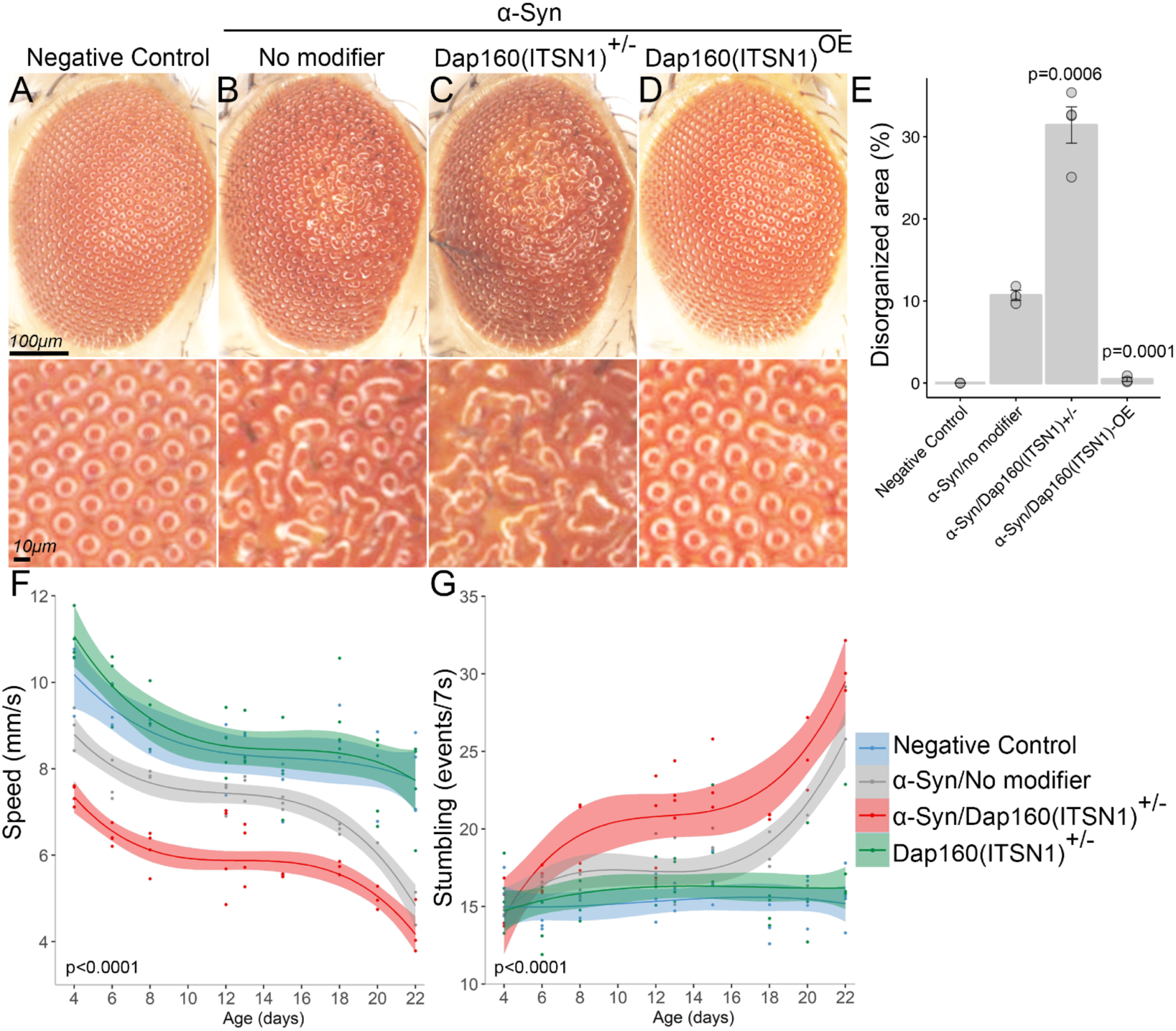
*Dap160(ITSN1)* modulates α-Syn-induced neurodegeneration in *Drosophila*. **(A-D)** Representative close-ups of *Drosophila* eyes, including the negative control (wild-type) **(A)**, α-Syn-expressing **(B)**, α-Syn/Dap160+/- **(C)**, and Dap160 overexpressing in the presence of α- Syn **(D)**. **(E)** Quantification of the disorganized eye area as a percentage of the total eye area for the genotypes shown in A-D. Error bars represent standard error of the mean. P-values were generated via ANOVA followed by Dunnet’s test using α-Syn/no modifier as control. (**F, G)** Motor performance as a function of age in the indicated genotypes. Two metrics are shown: speed **(F)** and stumbling events **(G)**. Lines represent spline fits; shaded areas are confidence intervals, p-values were obtained using a nonlinear random mixed effect model ANOVA applied to spline regressions of *α-Syn/no modifier* vs *α-Syn/ Dap160(ITSN1)^+/-^*. Complete genotypes are shown in **Table S12**. α-Syn = α-Synuclein; OE = overexpression.

We next studied the interaction of *Dap160* with α-Synuclein using a locomotor assay. Consistent with prior studies, neuronal expression of α-Synuclein in otherwise wild-type *Drosophila* led to a loss of climbing speed and increased stumbling events as the flies aged compared with age-matched controls (**Fig. 4F, G**). Heterozygous loss of *Dap160* worsened the phenotypes of α-Synuclein-expressing flies across all ages (**Fig. 4F, G**). Interestingly, *Dap160^+/-^* flies not expressing α-Synuclein displayed no eye or motor impairments, suggesting a synergistic interaction between α-Synuclein and *Dap160* haploinsufficiency. Altogether these results demonstrate a genetic interaction between *ITSN1* and α-Synuclein, further supporting the role of *ITSN1* haploinsufficiency in mediating PD risk.

## Discussion

To our knowledge, this study represents the largest sequencing-based case-control analysis of PD to date, including 3,702 cases and 233,378 controls in the discovery cohort from the UK Biobank and an additional 3,739 cases and 58,156 controls from two other independent cohorts (**Fig. 1**). Our variant-and gene-based association tests not only recapitulated multiple known associations, but also uncovered a novel association driven by ultra-rare PTVs in *ITSN1*. This association was unequivocally significant in a combined analysis across all three cohorts (**Fig. 2**). In the UK Biobank, *ITSN1* PTV carriers were at an over 10-fold increased risk of PD, ranking higher than every other detected risk locus. Our functional studies in *Drosophila* demonstrated that haploinsufficiency of the *ITSN1* homolog *Dap160* exacerbates compound eye degeneration and α-Synuclein-induced locomotor dysfunction (**Fig. 4**). Prior evidence has also shown that *Itsn1* null mice exhibit PD-related phenotypes, including abnormal gait, limb grasping deficits, and decreased prepulse inhibition.^25^ Collectively, these data establish *ITSN1* as a genetic cause of PD and suggest that it may interact with α-Synuclein-mediated pathogenic mechanisms.

There have been other recent studies focused on rare variant-driven associations with PD. Two recent studies^26,27^ focused on familial PD have independently converged on a rare variant in *RAB32* that was not sufficiently common in UKB to be included in our ExWAS. One of these studies^26^ included an exome-wide gene-level burden analysis but did not detect an association with *ITSN1.* One possible explanation for this is that they only ran a model that combined PTVs and missense variants, which may have obscured the PTV-specific effect. Another recent study performed a large, multi-cohort meta-analysis on rare protein-coding variation in 7,184 cases, 6,701 family-history-based proxy cases, and 51,650 controls.^5^ While this study suggested several new candidate PD genes, *ITSN1* was not among them. This could, in part, be attributed to the more homogenous and 4.5x larger control cohort in our discovery analysis. The diversity of findings across these studies underscores the importance of employing a range of approaches and leveraging large sample sizes to fully elucidate the complex genetic architecture of PD.

*ITSN1* encodes a multidomain scaffold protein that orchestrates protein-protein interactions during clathrin-mediated endocytosis and synaptic vesicle recycling.^14^ Notably, several other PD risk genes function in synaptic vesicle trafficking, including *SNCA, LRRK2, DNAJC6, SYNJ1, VPS35,* and *SH3GL2*.^28^ Dysfunction of this pathway has been implicated in multiple aspects of PD pathogenesis, such as the missorting and decreased degradation of alpha-synuclein and facilitating of its aggregation.^29^ Additionally, defects in synaptic vesicle endocytosis can disrupt the precise temporal coupling of exocytosis and endocytosis required for sustained neurotransmission, potentially contributing to synaptic dysfunction and neuronal loss in PD.^30^ Given its role as a scaffold protein, *ITSN1* haploinsufficiency may exacerbate these processes by destabilizing endocytic protein networks and impairing vesicle recycling. This is evidenced by decreased endocytosis rate and abnormal endosomes in *Itsn1* null mice,^31^ and reduced localization of critical endocytic proteins, enlarged vesicles, and aberrant synaptic formation in *Dap160* null *drosophila*.^24^ However, further studies are needed to elucidate the precise role of *ITSN1* in human PD pathogenesis and its potential as a therapeutic target.

*ITSN1* haploinsufficiency has also recently been implicated in moderate forms of ASD.^13^ Other studies have also highlighted a convergence between ASD and PD, with preliminary data suggesting that people with ASD are three times more likely to develop parkinsonism.^32^ Additionally, 22q11.2 deletion syndrome, which can cause ASD, has emerged as a genetic risk factor for early-onset PD.^33^ Interestingly, the association between *ITSN1* and ASD was driven in part by rare PTVs that probands inherited from unaffected parents. The spectrum of phenotypes associated with *ITSN1* haploinsufficiency, ranging from apparently unaffected carriers to individuals with increased risk of childhood-onset ASD or adult-onset PD, suggests variable penetrance and/or expressivity of *ITSN1* mutations. This genetic variability might lead to different phenotypic outcomes depending on additional genetic background, environmental exposures, or epigenetic modifications. Future studies aimed at identifying these modifiers would be critical for prognostication and could also unveil potential therapeutic avenues for mitigating *ITSN1*-mediated clinical disease.

Our study also has limitations. Foremost, small sample sizes limited our ability to detect associations among individual non-European populations, and there is clear evidence that population-specific variation can exert substantial risk for PD.^34^ It is critical that we continue expanding sequencing efforts to improve the representation of non-European ancestries, both for health equity and for fully elucidating the genetic architecture of human disease. Additional studies in rodent models and human cellular models are critical to elucidate the complex biological mechanisms of *ITSN1-*related PD.

Overall, our study leveraged population-scale whole-genome sequencing data to uncover a novel association between ultra-rare protein-truncating variants in *ITSN1* and a substantially increased risk of PD. Our functional studies provide critical evidence supporting ITSN1’s role in PD pathogenesis and suggest a potential interaction with α-Synuclein-mediated neurotoxicity. These findings add to the growing body of evidence implicating synaptic vesicle trafficking defects in PD and highlight ITSN1 as a potential therapeutic target. Importantly, this study also underscores the immense value of large-scale sequencing efforts for identifying rare variant contributions to complex diseases like PD.

## Methods

### Genetic association analysis

#### Samples

To permit binary genetic association analysis, we defined cohorts of people with PD (cases) and people without any neurological disorder (controls) from several datasets (see **Table 2**).^35–39^ Samples retained for analysis were determined by per-dataset sample quality control (QC) protocols (Supp. Methods) and assigned to inferred super-population ancestry cohorts. To control for sex as a possible confounder in the discovery analyses (see Supp. Methods), the QC protocol includes a sex-rebalancing step to address instances of unequal male:female ratio across cases and controls by downsampling the sex overrepresented in the control cohort to match the ratio in cases. Age of PD diagnosis was ascertained for UKB cases as the interval between date of birth (UKB field 33) and of first occurrence for any code mapped to ICD-10 G20 (UKBfield 131022), using the R *lubridate* package (v1.9.2).

**Table 2.** Summary of case and control cohort definitions for genetic association analysis across datasets. .*Cases from AMP-PD were combined with controls from All of Us and analyzed as a combined cohort.

| Dataset<br>(reference) | Definition |  |
| --- | --- | --- |
|  | Cases | Controls |
| UK Biobank<sup>35</sup> | Presence of ICD-10 G20 code recorded across UK Biobank fields 41270, 41202, 40001, 40002, 40006, 20002, 42031, 42033, and 131023 | Absence of any ICD-10 G-chapter code in medical history, indicating diagnosis for a disease of the nervous system (see previous study)<sup>6</sup> |
| AMP-PD<sup>*36</sup> | Labelled as 'Parkinson's disease' or 'Idiopathic PD' by site investigator | N/A |
| All of Us<sup>*37</sup> | N/A | No diagnosis of a mental, behavioral, and neurodevelopmental disorder (F01-F99 ICD-10-CM code) and no diagnosis of a disease of the nervous system (G00-G99 ICD-10-CM code) |
| 100,000 Genomes Project<sup>38,39</sup> | Recruitment for "Early onset and familial Parkinson's Disease" OR presence of ICD-10 G20 code in hospital records | Not relative of proband recruited for "Neurology and neurodevelopmental disorders" |

#### Statistical analysis

Variant-level genetic associations were tested under allelic (p allele versus q allele), dominant (pp versus pq or qq) and recessive (pp or pq versus qq) genetic models. Gene-level associations were tested using 10 collapsing analysis models (nine dominant and one recessive) which aggregate coding-sequence variation at the gene level. Dominant collapsing models compare people with at least one QV in a gene versus people without a QV. The recessive model compares people either homozygous for a QV or with at least two distinct QVs (assumed to be compound heterozygous) in a gene against those with one or fewer QVs. QVs are determined by model-specific parameters (**Table S3**) and model-agnostic variant QC filters; see Supp. Methods for details on Variant QC across variant-and gene-level analyses.

A two-sided Fisher’s Exact Test (FET) was used to test genetic associations to PD in UKB EUR across all variant-and gene-level models using the internal AstraZeneca computational environment, Amazon Web Services Cloud Adoption Framework. Variants from the ExWAS analysis with MAF ≥0.001 and associations at p<1x10^-4^ were clumped into independent loci using plink2 (v5.10 final)^40^ under default parameters (’--clump-r2 0.5’ and ’-- clump-kb 250’), based on LD in a random subsample of 10,000 individuals from UKB EUR. A two-sided exact CMH test was used for pan-ancestry gene-level UKB analyses, stratified across four ancestry cohorts. CMH was implemented using the R *stats* package (v4.1.0) *mantelhaen.test*. All remaining two-sided FETs used for follow-up analyses were performed using the R stats package *fisher.test* function under default settings.

The *ptv* collapsing model was reimplemented in additional cohorts for replication of the gene-level *ITSN1* association (Supp. Methods). Per-cohort associations were tested in R as above using two-sided FET and two-sided CMH was used for cohort-stratified analysis.

Analytical robustness was assessed firstly based on genomic inflation, whereby lambda was estimated using a regression-based approach across p-values observed under each model and expected from an n-of-1 permutation of case-control labels to attain an empirical null distribution.^10^ In gene-level analysis we additionally evaluated results from an empirical-negative synonymous variant model.^10^

Following previous studies, discovery analyses were interpreted against a conservative significance threshold of p<1x10^-8^.^41^ This was deemed sufficient based on: the Bonferroni-corrected p-value thresholds of 2.9x10^-7^ (0.05/170,370= 0.05/(9_non-synonymous models_*18,930_tests_)) in gene-level and 6.1x10^-9^ (0.05/8,253,351= 0.05/(3_genetic models_*2,751,117_tests_)) for variant-level analysis; evaluation of the smallest p-values from the ‘n-of-1’ permutation (gene-level = 3.5x10^-6^, variant-level = 5.4x10^-7^) tests and from the synonymous (4.2x10^-5^) collapsing analyses. A lenient threshold of p<1x10^-6^ was used to capture sub-significant signals that may warrant follow-up. Replication analyses were conducted with a nominal significance threshold of p<0.05.

We have received an exception to the Data and Statistics Dissemination Policy from the *All of Us* Resource Access Board to demonstrate the results of our collapsing analysis using All of Us short-read WGS data.

### Network analysis

The top 20 ITSN1 interactors and the edges connecting them to known PD genes were identified and mapped using StringDB V.12.0. A two-tailed Fisher’s exact test was used to test for enrichment in PD gene interactions in the *ITSN1* network comparing: 1-the proportion of genes in the entire StringDB network that are first degree neighbors of the indicated PD genes (expected number of interactors) to 2-the proportion of genes in the *ITSN1* network that are first degree neighbors of the selected PD genes (observed number of interactors). *ITSN1* in the context of this comparison was not considered a PD gene, as to not artificially bias the result. Network was designed and visualized using Cytoscape V3.10.2.

### *In vivo* analysis

#### Drosophila strains

All strains were obtained from the Bloomington Drosophila Stock Center (https://bdsc.indiana.edu/Home/Search). To express human wild-type codon optimized α- Synuclein, we used BDSC UAS line 51375 (*P{w[+mC]=UAS-SNCA.J}1*) in combination with the pan-neuronal post mitotic driver *elav-GAL4(C155)* (*P{w[+mW.hs]=GawB}elav[C155],* BDSC-458) or the eye driver *GMR-GAL4* (*P{GAL4-ninaE.GMR}12*, BDSC-1104). To modulate expression of *Dap160*, the *Drosophila* homolog of *ITSN1*, we used the amorphic mutant lines *Dap160Δ1* (BDSC-24877) and *Dap160Δ2* (P*{w[+mC]=UAS-Dap160.K}4.1*, BDSC-24878), and the *UAS-Dap160* overexpression line BSDC-42695. Specific genotypes are shown in Table S12.

#### Drosophila motor performance

Age-dependent motor performance was assessed using ten age-matched females per-replica per-genotype as previously described.^42,43^ Females were collected over a 24-hour period and transferred into new media daily. Animals were kept at 25°C. Using an automated platform, flies were tapped to the bottom of a plastic vial and recorded for 7.5s as they climb back up. Videos were analyzed using custom software to assess the speed and stumbling events of each animal. Four trials were performed each day per replicate, with four replicates per genotype. Data were analyzed using a nonlinear random mixed effect model ANOVA applied to spline regressions, adjusting p-values for multiplicity using Holm’s procedure. All graphing and statistical analyses were performed in R. Negative controls with baseline disease and motor performance were established using a neutral UAS line.

#### Drosophila eye images

Fruit flies were raised at 29°C. Eye images were captured from adult (1-day old) female flies using the Leica MZ16 imaging system. Areas of the eye showing fused or missing facets were selected with ImageJ. The average ratio of “disorganized eye area/total eye area” was calculated for α-Synuclein-expressing fly genotypes. Data was analyzed using ANOVA followed by Dunnet’s post hoc test comparing to the *α-Syn/no modifier* genotype.

## Data Availability

Summary statistics for the genetic association tests are provided in Supplementary Tables. UK Biobank whole-genome data may be requested via application to the UK Biobank. Additional information about registration for access to the data is available at http://www.ukbiobank.ac.uk/register-apply/. Phenome-wide association study summary statistics are available through our PheWAS portal (http://azphewas.com/). We also used data from 100KGP (https://www.genomicsengland.co.uk/research/academic), AMP-PD (https://amp-pd.org), and All of Us (https://www.researchallofus.org/data-tools/workbench/). We also accessed MTR (http://mtr-viewer.mdhs.unimelb.edu.au) and gnomAD (https://gnomad.broadinstitute.org).

## Acknowledgements

We thank the participants and investigators in the UKB study who made this work possible (Resource Application Numbers 26041; 68601); the UKB Genome Sequencing Consortium which includes AstraZeneca, Amgen, GlaxoSmithKline, Johnson & Johnson, Wellcome Trust Sanger, UK Research and Innovation, and the UKB; the AstraZeneca Centre for Genomics Research Analytics and Informatics team for processing and analysis of sequencing data.

This research was made possible through access to data in the National Genomic Research Library (100kGP), which is managed by Genomics England Limited (a wholly owned company of the Department of Health and Social Care). The National Genomic Research Library holds data provided by patients and collected by the NHS as part of their care and data collected as part of their participation in research. The National Genomic Research Library is funded by the National Institute for Health Research and NHS England. The Wellcome Trust, Cancer Research UK and the Medical Research Council have also funded research infrastructure.

Data used in the preparation of this article were obtained from the Accelerating Medicine Partnership® (AMP®) Parkinson’s Disease (AMP PD) Knowledge Platform. For up-to-date information on the study, visit https://www.amp-pd.org. The AMP® PD program is a public-private partnership managed by the Foundation for the National Institutes of Health and funded by the National Institute of Neurological Disorders and Stroke (NINDS) in partnership with the Aligning Science Across Parkinson’s (ASAP) initiative; Celgene Corporation, a subsidiary of Bristol-Myers Squibb Company; GlaxoSmithKline plc (GSK); The Michael J. Fox Foundation for Parkinson’s Research; AbbVie Inc.; Pfizer Inc.; Sanofi US Services Inc.; and Verily Life Sciences. ACCELERATING MEDICINES PARTNERSHIP and AMP are registered service marks of the U.S. Department of Health and Human Services.

Clinical data and biosamples used in preparation of this article were obtained from the (i) Michael J. Fox Foundation for Parkinson’s Research (MJFF) and National Institutes of Neurological Disorders and Stroke (NINDS) BioFIND study, (ii) Harvard Biomarkers Study (HBS), (iii) National Institute on Aging (NIA) International Lewy Body Dementia Genetics Consortium Genome Sequencing in Lewy Body Dementia Case-control Cohort (LBD), (iv) MJFF LRRK2 Cohort Consortium (LCC), (v) NINDS Parkinson’s Disease Biomarkers Program (PDBP), (vi) MJFF Parkinson’s Progression Markers Initiative (PPMI), and (vii) NINDS Study of Isradipine as a Disease-modifying Agent in Subjects With Early Parkinson Disease, Phase 3 (STEADY-PD3) and (viii) the NINDS Study of Urate Elevation in Parkinson’s Disease, Phase 3 (SURE-PD3).

BioFIND is sponsored by The Michael J. Fox Foundation for Parkinson’s Research (MJFF) with support from the National Institute for Neurological Disorders and Stroke (NINDS). The BioFIND Investigators have not participated in reviewing the data analysis or content of the manuscript. For up-to-date information on the study, visit michaeljfox.org/biofind.. Genome sequence data for the Lewy body dementia case-control cohort were generated at the Intramural Research Program of the U.S. National Institutes of Health. The study was supported in part by the National Institute on Aging (program #: 1ZIAAG000935) and the National Institute of Neurological Disorders and Stroke (program #: 1ZIANS003154).

The Harvard Biomarker Study (HBS) is a collaboration of HBS investigators [full list of HBS investigators found at https://www.bwhparkinsoncenter.org/biobank/] and funded through philanthropy and NIH and Non-NIH funding sources. The HBS Investigators have not participated in reviewing the data analysis or content of the manuscript.

Data used in preparation of this article were obtained from The Michael J. Fox Foundation sponsored LRRK2 Cohort Consortium (LCC). The LCC Investigators have not participated in reviewing the data analysis or content of the manuscript. For up-to-date information on the study, visit https://www.michaeljfox.org/biospecimens).

PPMI is sponsored by The Michael J. Fox Foundation for Parkinson’s Research and supported by a consortium of scientific partners: 4D Pharma, Abbvie, AcureX, Allergan, Amathus Therapeutics, Aligning Science Across Parkinson’s, AskBio, Avid Radiopharmaceuticals, BIAL, BioArctic, Biogen, Biohaven, BioLegend, BlueRock Therapeutics, Bristol-Myers Squibb, Calico Labs, Capsida Biotherapeutics, Celgene, Cerevel Therapeutics, Coave Therapeutics, DaCapo Brainscience, Denali, Edmond J. Safra Foundation, Eli Lilly, Gain Therapeutics, GE HealthCare, Genentech, GSK, Golub Capital, Handl Therapeutics, Insitro, Jazz Pharmaceuticals, Johnson & Johnson Innovative Medicine, Lundbeck, Merck, Meso Scale Discovery, Mission Therapeutics, Neurocrine Biosciences, Neuron23, Neuropore, Pfizer, Piramal, Prevail Therapeutics, Roche, Sanofi, Servier, Sun Pharma Advanced Research Company, Takeda, Teva, UCB, Vanqua Bio, Verily, Voyager Therapeutics, the Weston Family Foundation and Yumanity Therapeutics. The PPMI investigators have not participated in reviewing the data analysis or content of the manuscript. For up-to-date information on the study, visit www.ppmi-info.org.

The Parkinson’s Disease Biomarker Program (PDBP) consortium is supported by the National Institute of Neurological Disorders and Stroke (NINDS) at the National Institutes of Health. A full list of PDBP investigators can be found at https://pdbp.ninds.nih.gov/policy. The PDBP investigators have not participated in reviewing the data analysis or content of the manuscript.

The Study of Isradipine as a Disease-modifying Agent in Subjects With Early Parkinson Disease, Phase 3 (STEADY-PD3) is funded by the National Institute of Neurological Disorders and Stroke (NINDS) at the National Institutes of Health with support from The Michael J. Fox Foundation and the Parkinson Study Group. For additional study information, visit https://clinicaltrials.gov/ct2/show/study/NCT02168842. The STEADY-PD3 investigators have not participated in reviewing the data analysis or content of the manuscript.

The Study of Urate Elevation in Parkinson’s Disease, Phase 3 (SURE-PD3) is funded by the National Institute of Neurological Disorders and Stroke (NINDS) at the National Institutes of Health with support from The Michael J. Fox Foundation and the Parkinson Study Group. For additional study information, visit https://clinicaltrials.gov/ct2/show/NCT02642393. The SURE-PD3 investigators have not participated in reviewing the data analysis or content of the manuscript.

We gratefully acknowledge All of Us participants for their contributions, without whom this research would not have been possible. We also thank the National Institutes of Health’s <u>All of Us Research Program</u> for making available the participant data examined in this study.

The Genotype-Tissue Expression (GTEx) Project was supported by the Common Fund of the Office of the Director of the National Institutes of Health, and by NCI, NHGRI, NHLBI, NIDA, NIMH, and NINDS. The data used for this manuscript were obtained from the GTEx Portal on 16/05/2024.

