## Supplementary Information for "Haploinsufficiency of *ITSN1* is associated with Parkinson’s disease"

### Supplementary Methods

#### UK Biobank dataset

##### *Sequencing and variant calling*

Whole-genome sequencing (WGS) data of the UKB participants were generated by deCODE Genetics and the Wellcome Trust Sanger Institute as part of a public-private partnership involving AstraZeneca, Amgen, GlaxoSmithKline, Johnson & Johnson, Wellcome Trust Sanger, UK Research and Innovation, and the UKB. The WGS sequencing methods have been previously described.[1,2] Briefly, genomic DNA underwent paired-end sequencing on Illumina NovaSeq6000 instruments with a read length of 2x151 and an average coverage of 32.5x. Conversion of sequencing data in BCL format to FASTQ format and the assignments of paired-end sequence reads to samples were based on 10-base barcodes, using bcl2fastq v2.19.0. Initial quality control was performed by deCODE and Wellcome Sanger, which included sex discordance, contamination, unresolved duplicate sequences, and discordance with microarray genotyping data checks.

UK Biobank genomes were processed at AstraZeneca using the provided CRAM format files. A custom-built Amazon Web Services cloud compute platform running Illumina DRAGEN Bio-IT Platform Germline Pipeline v3.7.8 was used to align the reads to the GRCh38 genome reference and to call small variants. Variants were annotated against the transcript for which they are most damaging using SnpEff v4.3[3] and Ensembl Build 38.92.[4]

##### *Sample QC*

Genome sequencing data from 490,560 people were downloaded from the UK Biobank in April 2023. Phenotypic data used to identify people with Parkinson's disease (cases) and non-neurological controls (see **Table 2**) were accessed in April 2022.

Sample-level filters were applied as previously described to derive a subset of UKB suitable for analysis.[5] We removed any person withdrawn from UKB and without linked WGS data. Where available, WGS data were checked for concordance with previous exome sequencing and genotyping array data releases using the KING relatedness software v2.2.3,[6] excluding people with <0.49 estimated kinship with their exome-sequencing or <0.465 with the array data.

Of the remaining samples, we retained only those with VerifyBamID FREEMIX[7] contamination <0.04 and ≥10x coverage across ≥94.5% of Consensus Coding Sequence (CCDS) database (release 22) bases.[8] The cohort was filtered further using the *ukb_gen_samples_to_remove* function from the ukbtools (v0.11.3)[9] R package to select the largest subset of individuals with ≤0.3536 kinship estimate from KING, removing any first-degree relatives.

The data were next stratified by inferred ancestry across four superpopulation cohorts: African (AFR), East Asian (EAS), European (EUR), and South Asian (SAS). Ancestry was inferred based on the 1000 Genomes phase 3 cohort[10] using the Peddy (v0.4.2) software package[11] and we retained people with ≥0.90 probability of belonging to an ancestry group. For EUR ancestry, which represents >90% of the dataset, we further restricted to people ≤4 standard deviations (SD) from the mean of the first four genetic principal components (PCs). To avoid analytical confounding, female controls were pseudo-randomly downsampled if there was a significant difference in the odds of being male across cases and controls from a given ancestry cohort (Fisher's Exact Test p-value <0.05). **Table S1** overviews per-ancestry cohorts before and after sex rebalancing, where applied.

##### *Variant-level (ExWAS) analysis*

The following in-sample variant QC metrics were applied to select variants for inclusion in the ExWAS analysis: coverage ≥10x; ≥0.20 of reads are for the alternate allele for heterozygous genotype calls; binomial test of alternate allele proportion departure from 50% in heterozygous state p $\geq 1 \times 10^{-6}$; GQ ≥20; Fisher Strand Bias (FS) ≤200 for indels and ≤60 for SNVs; root mean square mapping quality (MQ) ≥40; QUAL ≥30; read position rank sum score (RPRS) ≥−2; mapping quality rank score (MQRS) ≥−8; DRAGEN variant status = PASS; ≤10% of the cohort have missing genotypes. Additional out-of-sample QC filters were also imposed based on the gnomAD v2.1.1 exomes (GRCh38 liftover) dataset.[12] The site of all variants should have ≥10x coverage in ≥30% of gnomAD exomes and, if present, the variant should have an allele count ≥50% of the raw allele count (before removing low-quality genotypes) in the dataset. Variants with missing values on a particular filter were treated as passing that filter, and therefore retained unless failing another metric. Those present in fewer than 6 people from UKB EUR ancestry population, or failing QC in >20,000 people, were also removed.

##### *Gene-level collapsing analysis*

Ten collapsing models were defined to test gene-level associations with different types of coding-sequence variation, including a synonymous variant model as an empirical negative control. Per-model filters for selecting qualifying variants are described in **Table S3**. The following in-sample variant QC metrics were applied universally across the collapsing analysis models: coverage ≥10x; present in CCDS (release 22);[8] ≥0.8 of reads are for the alternate allele among homozygous genotype calls; [0.25,0.8] of reads are for the alternate allele for heterozygous genotype calls; binomial test of alternate allele proportion departure from 50% in heterozygous state p $\geq 1 \times 10^{-6}$; GQ ≥20; FS ≤200 for indels and ≤60 for SNVs; MQ ≥40; QUAL ≥30; RPRS ≥−2; MQRS ≥−8; DRAGEN variant status = PASS. Additional out-of-sample QC filters were also imposed based on the gnomAD v2.1.1 exomes (GRCh38 liftover) dataset.[12] For all variants, the site should have ≥10x coverage in ≥25% of gnomAD exomes. For variants present in gnomAD, we retained those with RPRS ≥-2 and MQ ≥30 in that cohort. Variants with missing values on a particular filter were treated as passing that filter and retained unless failing another metric.

#### AMP-PD dataset

##### *Sequencing and variant calling*

WGS data for cohorts contributing to AMP-PD were generated by Macrogen and the Uniformed Services University of Health Sciences using the Illumina HiSeq X Ten sequencer with samples coming from whole blood. Data processing was performed on the Google Cloud Platform and in Terra (https://app.terra.bio/). All data processing was performed against Build 38 of the Human Genome reference (GRCh38DH, 1000 Genomes Project version). Analysis was based on the AMP-PD joint-genotyping v4 release which consists of samples from the BioFIND, HBS, LBD, LCC, PDBP, PPMI, Steady PD, and SURE PD cohorts. These data were accessed in April 2024.

Single sample alignment and variant calling was performed using the Broad Institute's Functional Equivalent Workflow (https://github.com/amp-pd/amp-pd-workflows/tree/master/wgs/wgs-germline-snps-indels). Joint genotyping was performed using the Broad Institute's joint discovery workflow (https://github.com/amp-pd/amp-pd-workflows/tree/master/wgs/joint-discovery). Joint-genotyped VCFs were processed through the Variant Transforms tool from Google Cloud (https://github.com/googlegenomics/gcp-variant-transforms), annotating variants against multiple transcripts with the Ensembl Variant Effect Predictor (VEP)[13] and converting to Variant Transforms format.

##### *Sample QC*

AMP-PD WGS samples passing the following QC protocol, which has also been described previously,[14] were used in joint-genotyping. Samples were retained in accordance with the following inclusion criteria: VerifyBamID FREEMIX ≤0.03, Mean coverage ≥25 reads per variant, joint calling missingness <0.05, Transition transversion ratio (TiTv) ≥2 for variants in dbSNP (v138), deviation from expected autosomal homozygous genotype counts (Absolute method-of-moments F ≤0.15). Sex was inferred from genetic data using plink, and samples with an explicit mismatch to self-reported sex (‘male’ or ‘female’) were removed.

The KING relatedness software was used for estimation of relatedness in the cohort and to infer ancestry. Concordance between WGS and NeuroX genotyping array[15] samples was tested where available, excluding people with matched samples predicted to be less closely related than duplicates / MZ twins. A deduplication step was performed across the WGS samples, excluding the lower-quality WGS sample from each pair predicted to be duplicates / MZ twins.

Ancestry was inferred based on principal component analysis against the HapMap 3 reference panel.[16] Continental ancestries were assigned to people within ±6 standard deviations of the mean of PC1 and PC2 for that population in the HapMap reference. Conflicts between self-reported race and genetically-inferred ancestry were resolved by removing people inferred to be ‘African’ or ‘Asian’ and clinically-reported ‘white’ and those inferred to be ‘European’ and clinically-reported as ‘African’ or ’Asian’ from joint-genotyping. For the current study, we prioritized analysis of a homogenous European ancestry cohort and so retained only people reporting ‘white’ race that were not flagged as having admixed ancestry during the prior ancestry inference step.

##### *Gene-level analysis*

Analysis was performed based on gene-level aggregation of variants called during the central AMP-PD joint-genotyping protocol that passed GATK QC filters and occurring at sites present in CCDS (release 22).[8] The *ptv* collapsing analysis model was implemented in AMP-PD (see **Table S3**). Qualifying variants were included on the basis of consequences for any transcript of *ITSN1*.

#### The 100,000 Genomes Project (100kGP) dataset

##### *Sequencing and variant calling*

Whole-genome sequencing data were generated, as previously described, using TruSeq DNA polymerase-chain-reaction (PCR)–free sample preparation kit (Illumina) on the HiSeq2500 platform. Reads were aligned using the Isaac Genome Alignment Software, and the Platypus variant caller was used for small variant calling.[17] Variants were annotated using VEP (v105) with the gnomAD plugin included.[13]

##### *Sample QC*

Samples from 100kGP that were retained for analysis were determined in-line with the following QC protocol inclusion criteria: VerifyBamID FREEMIX contamination ≤0.03; aligned reads have ≥15x coverage across 95% of the genome with MQ >10; >90% concordance between variant calls from sequencing and matched genotyping array; median fragment size >250bp; <5% chimeric reads; >60% mapped reads; <10% AT dropout; concordance between self-reported and genetically determined sex.

Ancestry was inferred by training a random forest model upon PCs 1-8 from the 1000 Genomes phase 3 cohort.[10] People with >99% probability of European ancestry and <4SD from the mean of PCs 1-4 in the 100kGP inferred-European cohort were retained for analysis. Finally, we removed one from each pair of individuals estimated to have ≥2nd degree relatedness (kinship coefficient >0.0442) under the plink2 implementation of the KING-relatedness software. Here we followed a 3-step procedure, removing one from each pair of related cases with the *ukb_gen_samples_to_remove* R function, removing controls related to remaining cases, and finally using *ukb_gen_samples_to_remove* to removing one from each related pair of remaining controls. Sex-rebalancing was then performed as for the UK Biobank dataset so that the odds of being male was comparable in cases and controls.

##### *Gene-level analysis*

Variants within the 100kGP dataset that were retained for analysis passed the following in-sample inclusion criteria: site missingness ≤0.05; median coverage ≥10x; Median GQ ≥15; ≥0.25 heterozygous allele calls do not show significant ($p<0.01$) allele imbalance in binomial test; ≥0.50 complete genotype data for a variant; Hardy-Weinberg equilibrium mid $p>1x10^{-5}$ among unrelated samples of European ancestry.

We tested gene-level associations to *ITSN1* under the *ptv* model (see **Table S3**). Variants were mapped to consequences based on their impact upon the Matched Annotation from NCBI and EMBL-EBI (MANE) transcript for the gene (ENST00000381318).

#### All of Us dataset

The All of Us Research Program aims to obtain genetic and health data from one million or more people living in the United States to accelerate biomedical research and improve human health. So far, more than 740,000 people have joined the program. At the time that this study was conducted, All of Us contained short-read WGS data from 245,376 individuals, 123,063 of whom were classified as having European genetic ancestry (see below).

##### *Sequencing and variant calling*

Genome sequencing, variant discovery, sample QC and variant QC of All of Us data are explained in the recent study by the All of Us research program genomics investigators.[18] The materials explained here are obtained from that study and All of Us genomic quality report (https://support.researchallofus.org/hc/en-us/articles/4617899955092-All-of-Us-Genomic-Quality-Report).

To ensure accurate and consistent DNA sample extraction and sequencing, the All of Us Genome Centers and Biobank implemented standardized laboratory protocols, quality control methods, and validation experiments with known clinical samples and reference standards. PCR-free Barcoded WGS libraries were created using the Illumina Kapa HyperPrep kit. The libraries were then pooled and sequenced on the Illumina NovaSeq 6000 instrument. Following demultiplexing, initial QC analysis is conducted using the Illumina DRAGEN pipeline, which includes metrics at the lane, library, flow cell, barcode, and sample levels. This analysis also evaluates contamination, mapping quality, and concordance with genotyping array data processed separately from a different DNA aliquot. The Genome Centers utilize these metrics to assess if each sample meets program specifications. Once approved, the sequencing data is then sent to the Data and Research Center for additional quality control, joint calling, and distribution to the research community.

##### *Sample QC*

All of Us samples passing the following quality control metrics were used in joint calling and released to the research community. The quality control criteria include Mean coverage (threshold ≥30x), Genome coverage (threshold ≥90% at 20x, All of Us Hereditary Disease Risk gene (AoUHDR) coverage (threshold ≥95% at 20x), Aligned Q30 bases (threshold $\geq 8 \times 10^{10}$) and contamination ≤1%. Additional QC checks include fingerprint mismatch and array mismatch.

Ancestry was inferred across populations defined in gnomAD using a random forest classifier trained using a diverse reference dataset of 3,202 samples and 151,159 autosomal single-nucleotide polymorphisms. By projecting the All of Us samples onto the classifier's 16-dimensional PCA space, categorical ancestry predictions were generated. Since data from AoU were used as a non-neurological control cohort for analysis together with cases of European ancestry from AMP-PD, we restricted to those classified as having European ancestry. The dataset was then restricted to include only those with corresponding sex at birth and genetically-inferred sex; 119,089 samples remained.

AoU used the Hail pc_relate function to determine the kinship score to report any pairs with a kinship score >0.1. The kinship score is half of the fraction of the genetic material shared (ranges from 0.0 - 0.5); parent-child or siblings: 0.25, Identical twins: 0.5. We identified pairs of related individuals in the remaining cohort using the relatedness.tsv file provided by AoU, which lists all pairs of samples with a kinship score above 0.2. The first sample of each related pair was removed, leaving 114,415 unrelated individuals. A final dataset of 50,754 people remained after removing samples with any diagnosis under the ICD-10 neurological disease chapter (**Table 2**).

##### *Gene-level analysis*

AoU applied the following site-level quality control metrics on variants: GQ ≥20, DP ≥10, AB ≥ 0.2 for heterozygotes, ExcessHet <54.69 and QUAL ≥60 for SNPs and QUAL ≥69 for Indels. We restricted to variants in positions overlapping CCDS transcripts (release 22). Analysis was performed based on gene-level aggregation of variants with qualifying effects upon *ITSN1* based on the *ptv* model (see **Table S3**). Variants were annotated using SnpEff v4.3 and included on the basis of consequences for any transcript of *ITSN1*.

#### Power analysis for *ITSN1* replication

Power analysis was performed to assess the sample requirements for 80% power to replicate the association between PD and *ITSN1* PTVs at p=0.05. Assuming that qualifying ITSN1 PTVs occur in 0.0003 of people (**Table S8**, **Table S9**), we estimated that a total of ~17,500 cases and controls (at 1:1 sampling) would be required to detect an association with an odds ratio of 10, and ~25,000 for an odds ratio of 6 (**Fig. S8**).

### Supplementary Table Legends

**Table S1 - UK Biobank ancestry cohorts and sex-rebalancing statistics.**

Two-sided Fisher's Exact Tests were used to compare the difference in odds of being male versus female across cases and control cohorts of UK Biobank. In cohorts with a $p<0.05$, we downsampled the sex overrepresented in controls. The column 'N controls (male: female)' indicates the control cohort sample size before downsampling, and the column 'N rebalanced controls (male: female)' indicates control cohort sample size after downsampling, where performed.

**Table S2 - Variant-level association statistics across all genetic models for variants with associations with p<1x10-4**

Association statistics for allelic/dominant/recessive genetic models are shown for all variants, which are aligned to GRCh38 and indicated in the 'variantID' column in the format CHR-POS-REF-ALT (chromosome, position, reference allele, alternate allele). The columns 'Gene_Symbol', 'hgnc_id', and 'ensembl_gene_id' label the gene for which each variant is identified as most damaging using SnpEff (v4.3). Rows are sorted by p-value from smallest to largest, with corresponding odds ratios and 95% confidence intervals. Minor allele frequencies (MAF) are given overall and for cases and controls individually. Variants with overall MAF ≥0.001 were clumped into independent loci based on the most significant genetic model for each variant; those with MAF<0.001 were not entered into clumping; The 'clumping_info' column labels variants as 'index', 'clumped', or 'LowMAF_not_clumped'; For index variants, the 'clumping_nClumped' column indicates the number of variants clumped within that column. For clumped variants, the 'clumping_clumpedIntoVariant' column indicates the associated index variant; '†' marks in the 'annotation' column flag variants with associations at $p<1x10^{-6}$ that are situated near a large chromosome 17 inversion (GRCh38: chr17:45,495,836-46,707,123)[19] and which may be subject to long-spanning linkage disequilibrium not fully captured during clumping. The "Gene_known_in" column labels variants mapped to genes previously implicated in Parkinson's disease, Alzheimer's disease, or amyotrophic lateral sclerosis (ALS). The genotype counts for reference allele 'p' and alternate allele 'q' are given in the columns 'caseGenotypes' and 'controlGenotypes', using the format 'pp/pq/qq' for autosomal and pseudo-autosomal variants and 'pp/pq/qq/p/q' for X-chromosome variants not in the pseudo-autosomal region, representing hemizygous male genotypes separately. The columns 'caseRefCount', 'caseAltCount', 'ctrlRefCount', 'ctrlAltCount' indicate the cell counts for the contingency table analyzed using Fisher's Exact Test; counts are derived from the genotypes indicated in 'caseGenotypes' and 'controlGenotypes' and depend upon to the tested genetic model (see Methods).

**Table S3 - Qualifying variant models.**

* reflects the gnomAD global_raw minor allele frequency (MAF) unless otherwise specified

^ reflects the maximum percentage of UKB genome sequences permitted to either have ≤10x coverage at variant site or carry a low-confidence variant failing at least one of the quality-control thresholds applied for collapsing analysis. The threshold is set with varying stringency for the large-scale European ancestry (EUR) cohort according to the Internal MAF threshold for each model; a default threshold of ≤0.005% is set across all models for the smaller African (AFR), East Asian (EAS), and South Asian (SAS) ancestry cohorts.

† For some models, a more lenient Internal MAF threshold is used in replication analysis because the datasets are substantially smaller than UKB and the analysis specifically targets genes with evidence of association to PD from the initial discovery analyses.

Ø reflects the probability score for a variant from gnomAD's random forest filter; higher probability scores indicate higher confidence of a true allele.

Ω Variant consequences:
Synonymous: synonymous_variant.
PTV: exon_loss_variant, frameshift_variant, start_lost, stop_gained, stop_lost, splice_acceptor_variant, splice_donor_variant, gene_fusion, bidirectional_gene_fusion, rare_amino_acid_variant, transcript_ablation.
Missense: missense_variant&splice_region_variant, missense_variant.
Non-synonymous: exon_loss_variant, frameshift_variant, start_lost, stop_gained, stop_lost, splice_acceptor_variant, splice_donor_variant, gene_fusion, bidirectional_gene_fusion, rare_amino_acid_variant, transcript_ablation, conservative_inframe_deletion, conservative_inframe_insertion, disruptive_inframe_insertion, disruptive_inframe_deletion, missense_variant&splice_region_variant, missense_variant, protein_altering_variant

UKB = UK Biobank; 100kGP = 100,000 Genomes project

**Table S4 - Gene-level association statistics in UK Biobank European ancestry cohort across all models for genes with associations at $p<1x10^{-4}$.**
Rows are sorted by p-value from smallest to largest. The 'Gene_known_in' column labels whether each gene has been previously implicated in ALS, Alzheimer's disease, or Parkinson's disease.

**Table S5. Breakdown of *GBA1* variants included within the *flexnonsynmtr* collapsing model analysis of UK Biobank Europeans.**
Variants were annotated to consequences against the MANE *GBA1* transcript (ENST00000368373) using SnpEff v4.3t against genome build GRCH38.92. Two-tailed Fisher's exact tests were performed to identify variants most influential upon the *GBA1* gene *flexnonsynmtr* model association. The "Abs_log10_pValueChange" column indicates the magnitude of difference from the main association result ($p=2.41x10^{-9}$) after removing a given variant, letting Abs_log10_pValueChange=1 indicate 1 order of magnitude difference from the main result; rows of the table are sorted by Abs_log10_pValueChange from largest to smallest.

The columns “n_UKBEUR_cases” and “n_UKBEUR_ctrls” respectively indicate the number of cases and controls harboring each variant.

**Table S6 - Gene-level association statistics in UK Biobank Pan-ancestry cohort across all models for genes with associations at $p<1x10^{-4}$.**
Rows are sorted by p-value from smallest to largest. The 'Gene_known_in' column labels whether each gene has been previously implicated in ALS, Alzheimer's disease, or Parkinson's disease. Counts for "n controls without QV", "n controls with QV", "n cases without QV" and "n cases with QV" are provided for each of the ancestry groups analyzed: African (AFR), East Asian (EAS), European (EUR), and South Asian (SAS). QV = variant qualifying for collapsing model.

**Table S7 - Breakdown of ITSN1 variants present in each cohort.**
Variants are annotated to consequences against the MANE *ITSN1* transcript (ENST00000381318) and, for variants without a protein-truncating effect upon ENST00000381318, the protein-coding transcript for which they are most damaging. Annotation was performed using SnpEff v4.3t against genome build GRCH38.92. Rows of the table are sorted by variant position.

UKBEUR = UK Biobank European-ancestry cohort; 100kGP = 100,000 Genomes Project; AMP-PD = Accelerating Medicines Partnership - Parkinson's Disease; AoU = All of Us.

**Table S8 - Cumulative frequency of ITSN1 PTVs qualifying for *ptv* model across PD and control cohorts.**

**Table S9 - Summary of *ITSN1* variants present in gnomAD (v2.1.1 exomes) that pass ptv collapsing model filters.**
The cumulative variant frequency in Non-Finnish Europeans is 0.000313, the sum of AC_nfe/AN_nfe for each variant. For the non-Finnish European non-neurological control cohort the frequency is 0.000214, a sum of non_neuro_AC_nfe/non_neuro_AN_nfe.

**Table S10 - Transcript-specific analysis of *ITSN1 ptv* gene-level collapsing model in the UK Biobank European ancestry cohort.**

Variants qualifying for the main transcript-agnostic ITSN1 ptv model analysis were reannotated against specific ITSN1 transcripts using SnpEff v4.3t against genome build GRCH38.92. Variants were included in the analysis when they had a PTV effect upon that transcript (**Table S3**).

**Table S11 – Mantis-ML v2 results.**
Prioritization of Human Phenotype Ontology terms most likely to be associated with *ITSN1* by Mantis-ML v2

**Table S12 – *Drosophila* genotypes.**

Complete *Drosophila* genotypes for animals used in figure 4.

#### Supplementary Figures

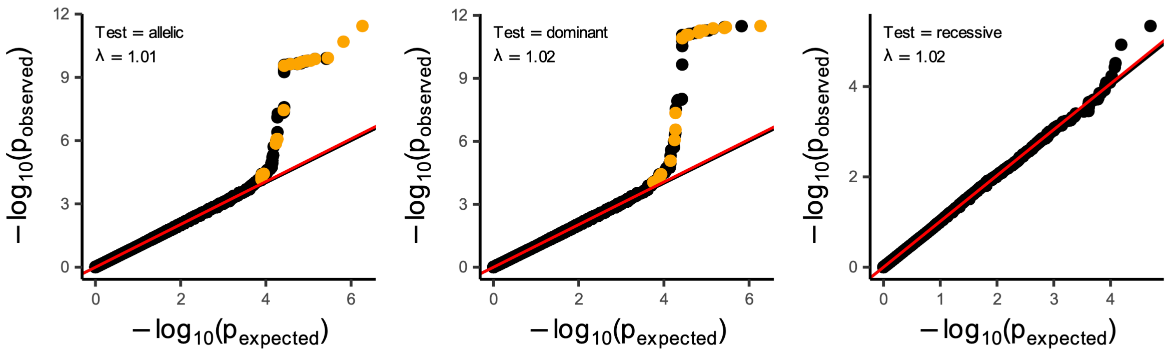

**Figure S1. QQ plots for observed and expected -log10(p) values in the variant-level analysis.** Orange point shading labels genes that have previously been associated to Parkinson's disease and have associations at $p<1x10^{-4}$. The null-distribution of expected p-values is defined based on n-of-1 permutation of phenotype group for each genetic test. The lambda (λ) genomic inflation factor was calculated under a regression-based approach.

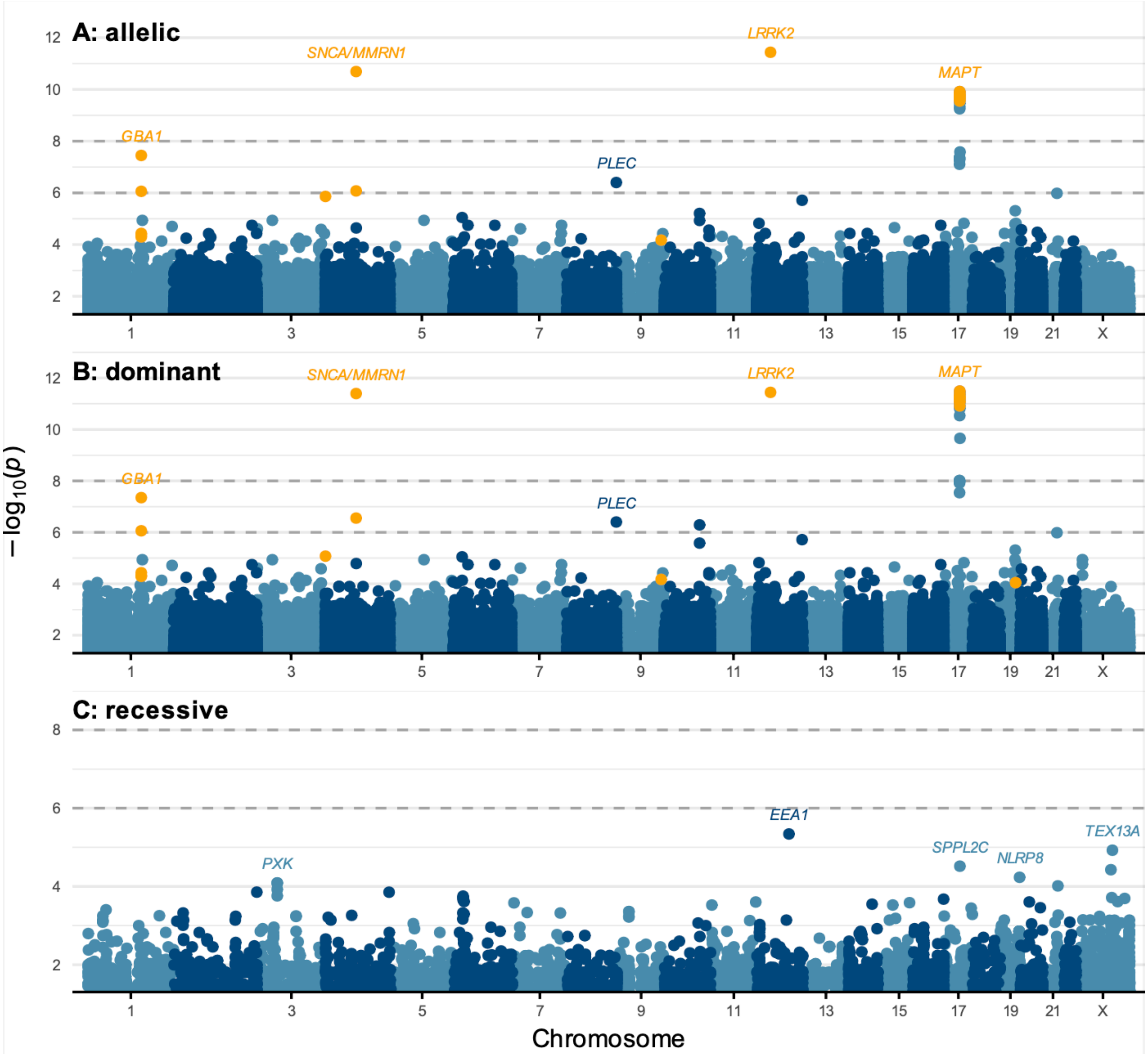

**Figure S2. Manhattan plots of variant-level associations to Parkinson's disease under each genetic model.** Orange point shading labels variants annotated against genes that have previously been associated to PD and have associations at $p<1x10^{-4}$; Horizontal dashed lines indicate the significance ($p<1x10^{-8}$) and suggestive ($p<1x10^{-6}$) thresholds for the analyses.

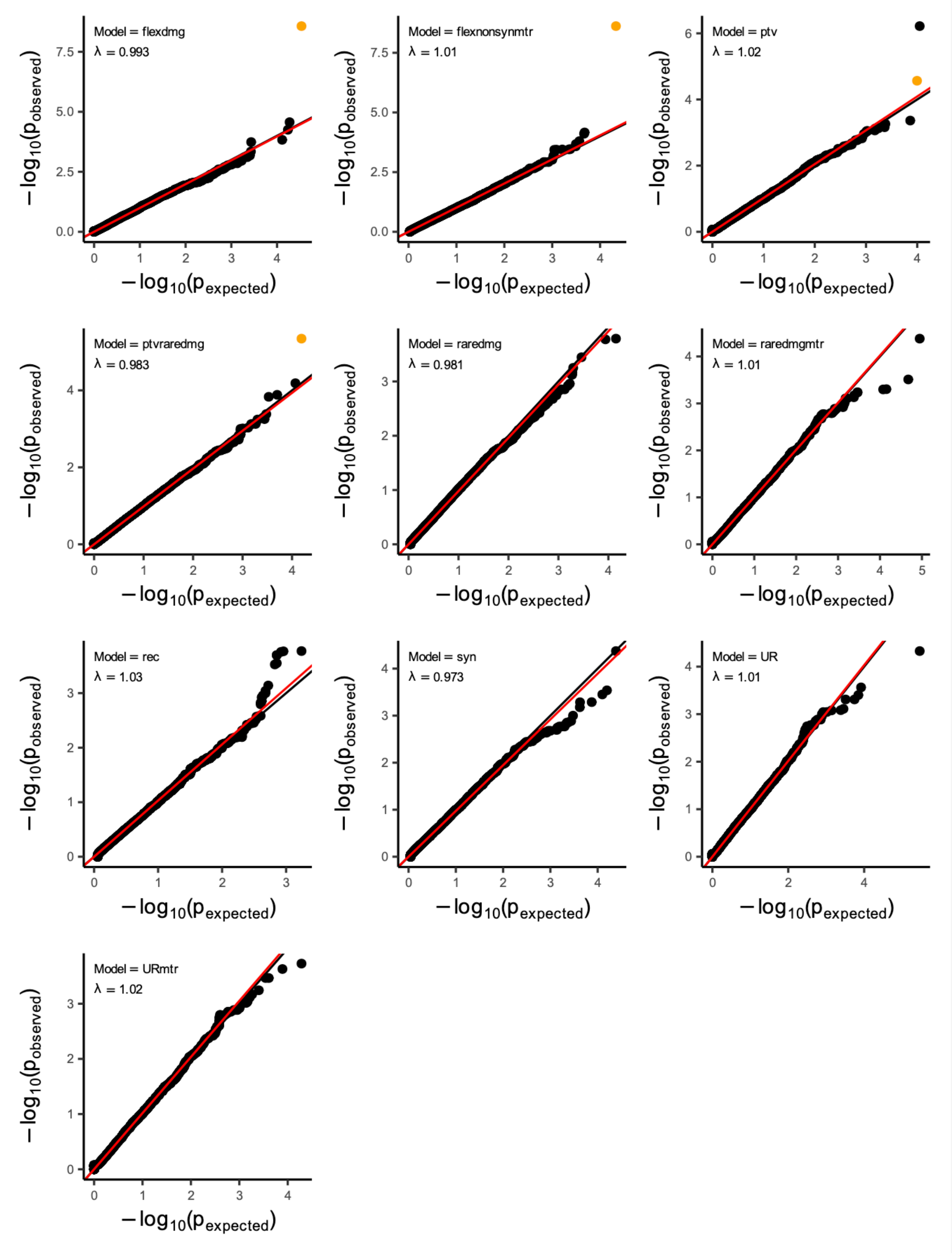

**Figure S3. QQ plots for observed and expected -log10(p) values in the gene-level analysis.** Orange point shading labels variants mapped to genes that have previously been associated to Parkinson's disease and have associations at $p<1x10^{-4}$. The null-distribution of expected p-values is defined based on n-of-1 permutation of phenotype group for each genetic test. The lambda (λ) genomic inflation factor was calculated under a regression-based approach.

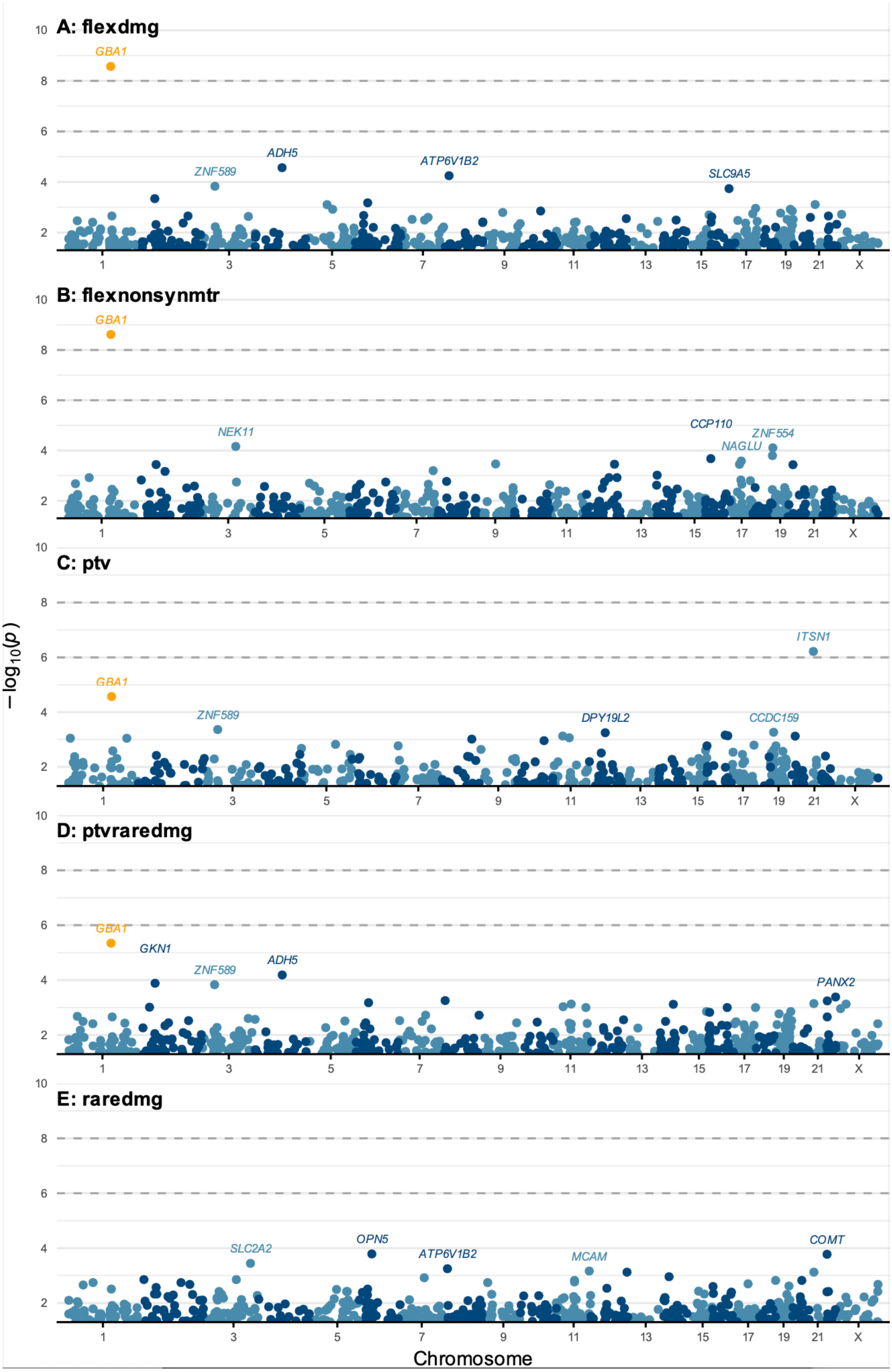

A: flexdmg
GBA1
ADH5
ZNF589
ATP6V1B2
SLC9A5
B: flexnonsynmtr
GBA1
NEK11
CCP110
ZNF554
NAGLU
C: ptv
ITSN1
GBA1
ZNF589
DPY19L2
CCDC159
D: ptvraredmg
GBA1
GKN1
ADH5
ZNF589
PANX2
E: raredmg
SLC2A2
OPN5
ATP6V1B2
MCAM
COMT
−log10(p)
Chromosome
1 3 5 7 9 11 13 15 17 19 21 X

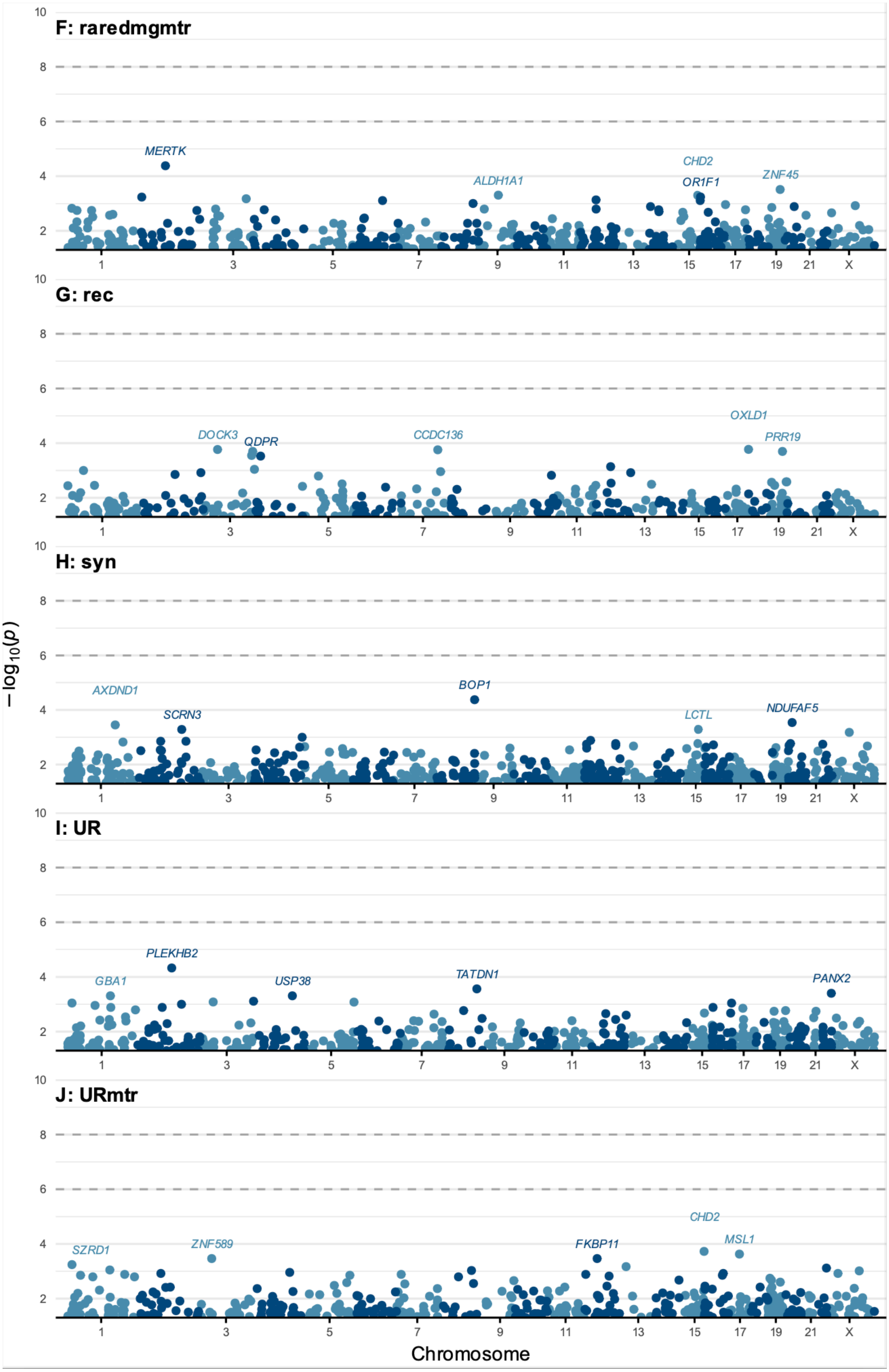
F: raredmgmtr
MERTK
ALDH1A1
CHD2
OR1F1
ZNF45
G: rec
DOCK3
QDPR
CCDC136
OXLD1
PRR19
H: syn
AXDND1
SCRN3
BOP1
LCTL
NDUFAF5
I: UR
GBA1
PLEKHB2
USP38
TATDN1
PANX2
J: URmtr
SZRD1
ZNF589
FKBP11
CHD2
MSL1
$-\log_{10}(p)$
Chromosome

**Figure S4. Manhattan plots of gene-level associations to Parkinson's disease under each collapsing model.** Orange point shading labels genes that have previously been associated to PD and have associations at $p<1x10^{-4}$; Horizontal dashed lines indicate the significance ($p<1x10^{-8}$) and suggestive ($p<1x10^{-6}$) thresholds for the analyses.

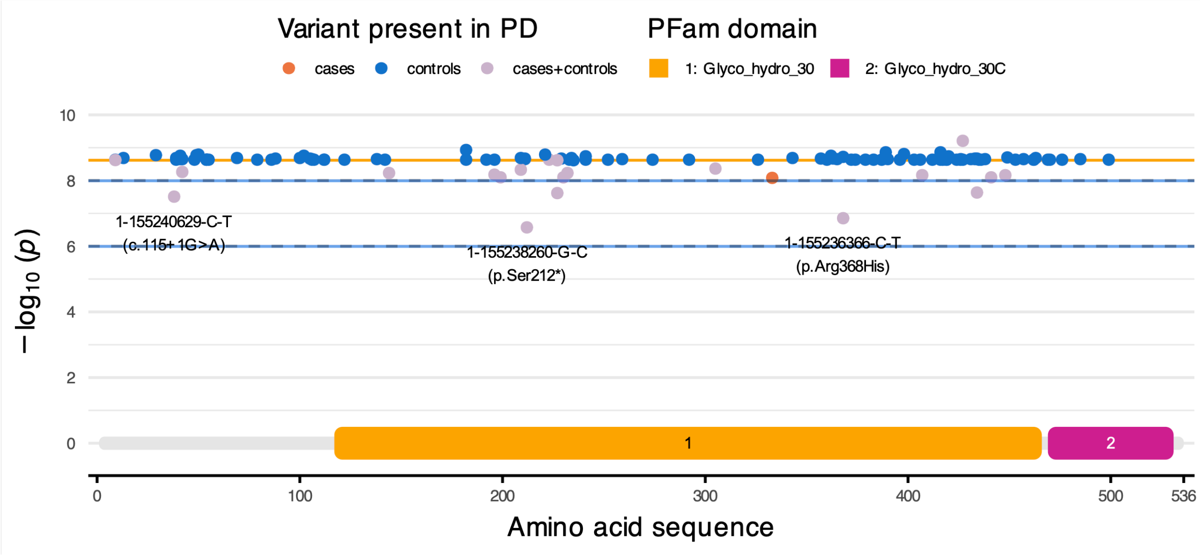

**Figure S5. Gene-level association results in UK Biobank European ancestry cohort 'leave-one-out' analyses.** Two-tailed Fisher's exact tests were performed for the *GBA1* gene under the *flexnonsynmtr c*ollapsing model after removal of a given qualifying variant. x-axis positions of points indicate the location of the removed variant relative to the amino acid sequence of the GBA1 MANE transcript (ENST00000368373) and PFam domains of the corresponding protein (P04062). The orange horizontal line on each panel indicates the association result from the main analysis ($p=2.41x10^{-9}$). Greater deviation from this line indicates greater influence of the removed variant upon the association result. Variants with >1 absolute difference in $-log_{10}(p)$ between the main and leave-one-out analysis results are labelled in the format 'variant ID (MANE transcript consequence: protein impact [or coding sequence impact for splice variants])'. Gray hatched lines indicate significance ($p<1x10^{-8}$) and suggestive ($p<1x10^{-6}$) thresholds for the study (Methods).

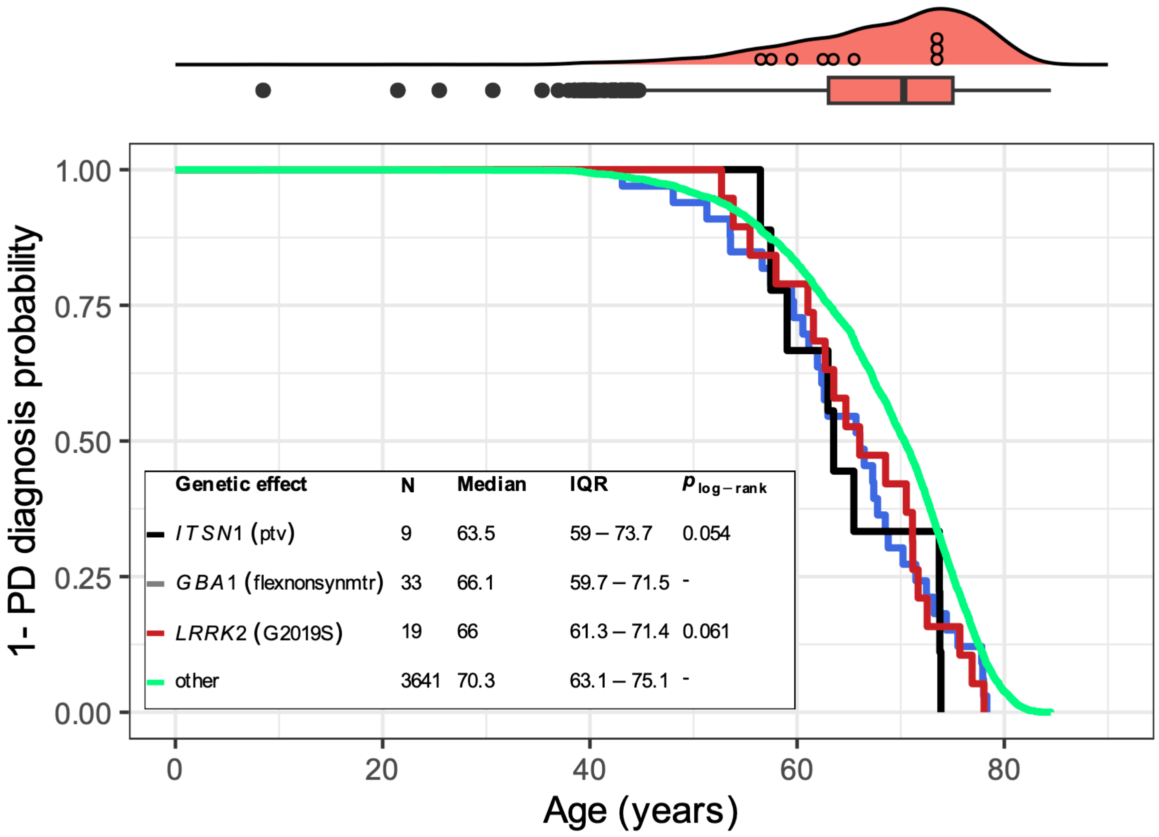

**Figure S6. Trends in age of Parkinson's disease diagnosis according to genetic susceptibility.** The density curve at the top of the figure and corresponding boxplot show the distribution in age of PD diagnosis across the UK Biobank European-ancestry cohort; black circular points within the density curve show age of diagnosis among people with a qualifying *ITSN1* PTV, binned by year. The Kaplan-Meier curve shows time from birth until age of PD diagnosis, stratifying by genetic effect; Trends in *ITSN1* and *GBA1* are shown for people harboring qualifying variants captured in the gene-level collapsing model most significant for each gene (see **Table 3**). Trends in *LRRK2* are shown for carriers of the well-established p.Gly2019Ser (12-40340400-G-A) variant; The 'other' group comprises the remaining PD cohort, without these genetic burdens. The inset table provides descriptive statistics for each group, and the '$p_{log\text{-}rank}$' values are for a log-rank test of the difference in survival for each group relative to the 'other' group; non-significant test results are not surprising since specific genetic effects occur in so few people.

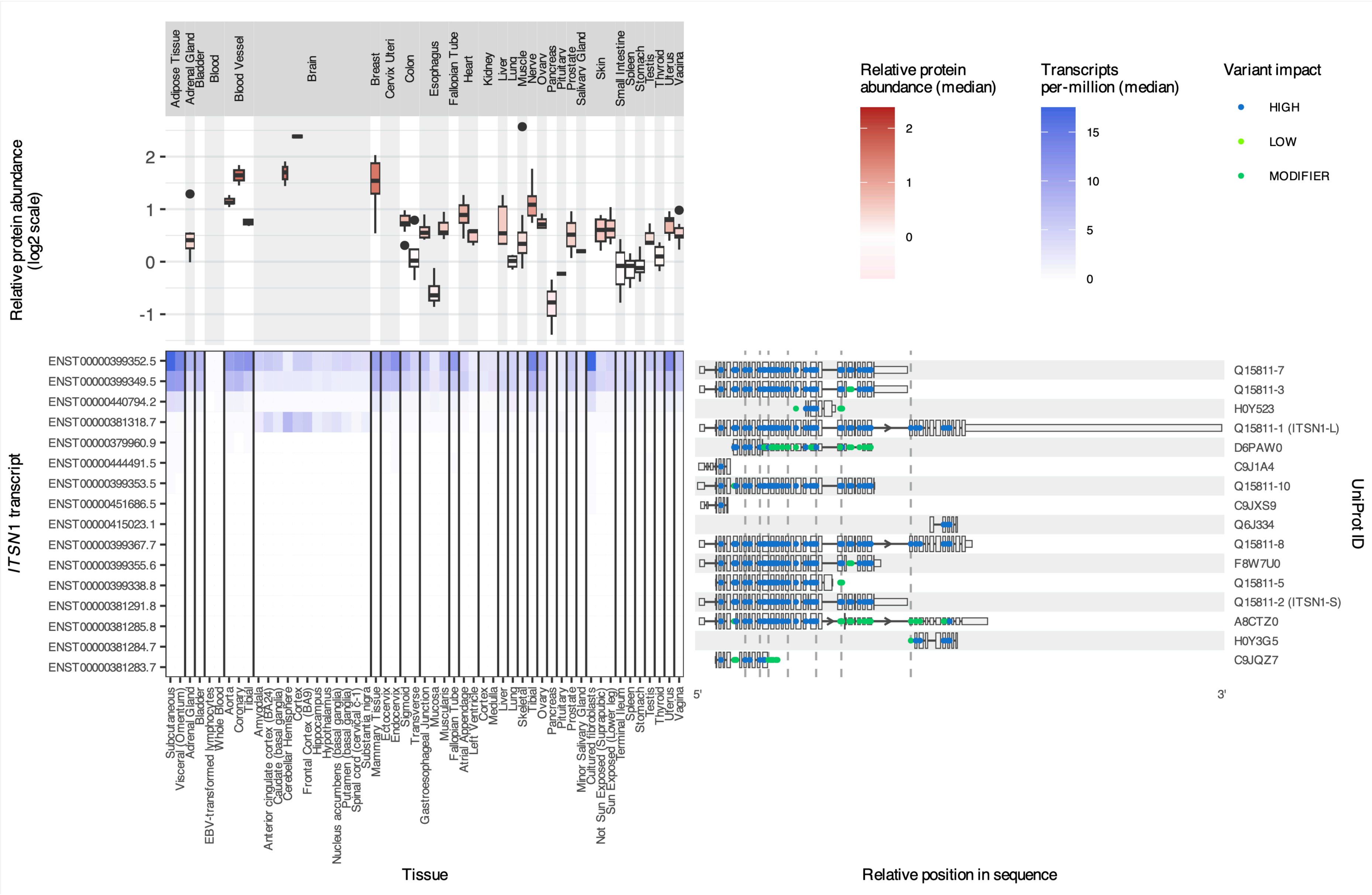

Adipose Tissue
Adrenal Gland
Bladder
Blood
Blood Vessel
Brain
Breast
Cervix Uteri
Colon
Esophagus
Fallopian Tube
Heart
Kidney
Liver
Lung
Muscle
Nerve
Ovary
Pancreas
Pituitary
Prostate
Salivary Gland
Skin
Small Intestine
Spleen
Stomach
Testis
Thyroid
Uterus
Vagina
Relative protein abundance (log2 scale)
Relative protein abundance (median)
Transcripts per-million (median)
Variant impact
HIGH
LOW
MODIFIER
ITSN1 transcript
ENST00000399352.5
ENST00000399349.5
ENST00000440794.2
ENST00000381318.7
ENST00000379960.9
ENST00000444491.5
ENST00000399353.5
ENST00000451686.5
ENST00000415023.1
ENST00000399367.7
ENST00000399355.6
ENST00000399338.8
ENST00000381291.8
ENST00000381285.8
ENST00000381284.7
ENST00000381283.7
Q15811-7
Q15811-3
H0Y523
Q15811-1 (ITSN1-L)
D6PAW0
C9J1A4
Q15811-10
C9JXS9
Q6J334
Q15811-8
F8W7U0
Q15811-5
Q15811-2 (ITSN1-S)
A8CTZ0
H0Y3G5
C9JQZ7
UniProt ID
Subcutaneous
Visceral (Omentum)
Adrenal Gland
Bladder
EBV-transformed lymphocytes
Whole Blood
Aorta
Coronary
Tibial
Amygdala
Anterior cingulate cortex (BA24)
Caudate (basal ganglia)
Cerebellar Hemisphere
Cortex
Frontal Cortex (BA9)
Hippocampus
Hypothalamus
Nucleus accumbens (basal ganglia)
Putamen (basal ganglia)
Spinal cord (cervical c-1)
Substantia nigra
Mammary Tissue
Ectocervix
Endocervix
Sigmoid
Transverse
Gastroesophageal Junction
Mucosa
Muscularis
Fallopian Tube
Atrial Appendage
Left Ventricle
Cortex
Medulla
Liver
Lung
Skeletal
Tibial
Ovary
Pancreas
Pituitary
Prostate
Minor Salivary Gland
Cultured fibroblasts
Not Sun Exposed (Suprapubic)
Sun Exposed (Lower leg)
Terminal Ileum
Spleen
Stomach
Testis
Thyroid
Uterus
Vagina
Tissue
5'
3'
Relative position in sequence

**Figure S7. *ITSN1* PTVs impact multiple protein-coding transcripts.** The heatmap displays expression of different protein-coding *ITSN1* transcripts across tissue samples from GTEx, ordered from most to least highly expressed. The panel to the right illustrates the sequence of each transcript and colored points show the location and impact of each *ITSN1* variant from the *ptv* model collapsing analysis against each transcript; HIGH impact indicates a protein-truncating effect; The positions of qualifying PTVs present in people with Parkinson's disease are marked by vertical hatched lines across this panel; intronic sequences that do not overlap exons of another transcript have been shortened to better visualize exons; bars for 5' and 3' untranslated regions are half the height of bars showing the coding sequence. The boxplots above the heatmap display estimates of relative ITSN1 protein abundance across 32 tissue sites in a subset of the GTEx cohort.[20]

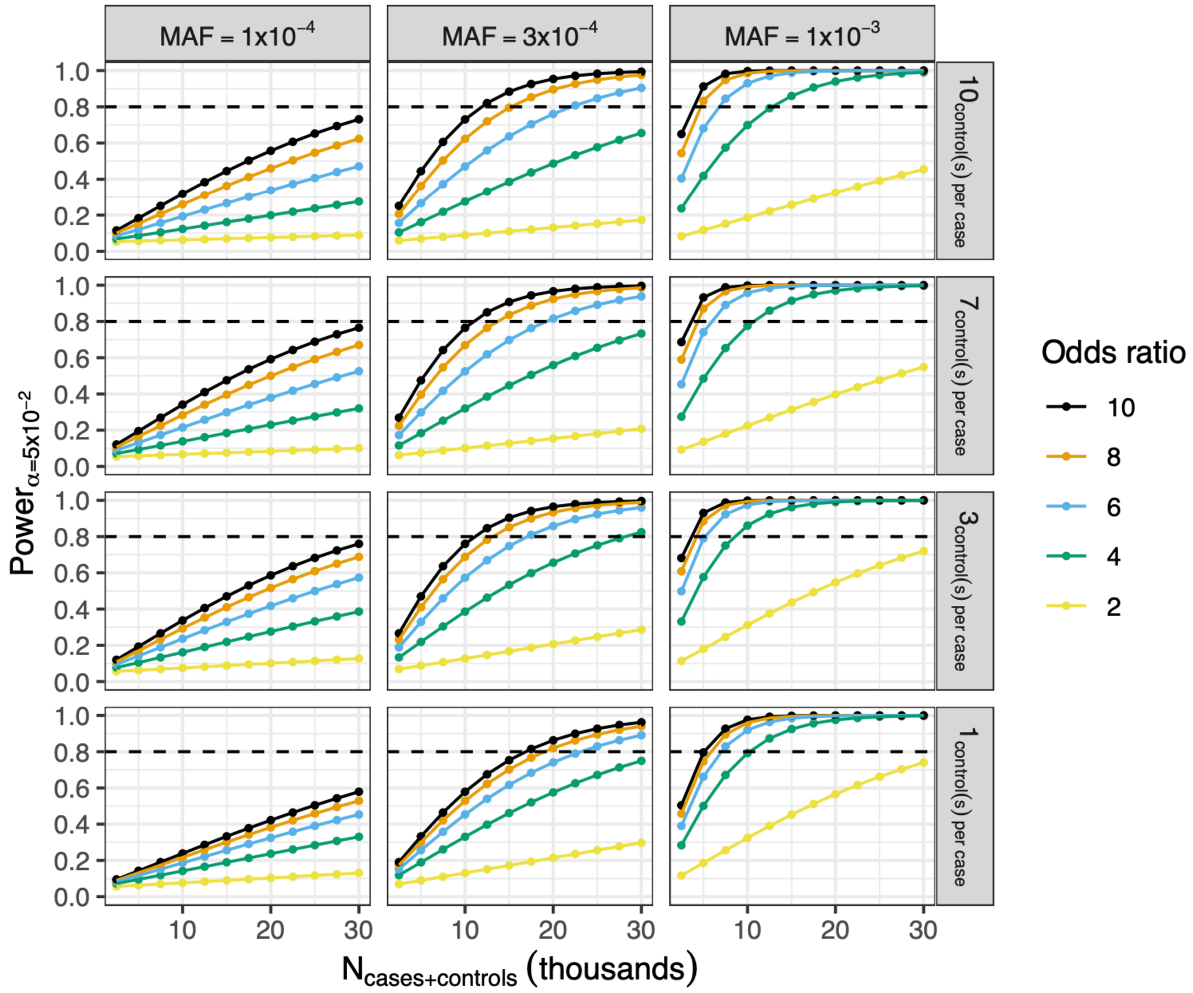

**Figure S8. Analysis of power to detect a dominant genetic association for a rare variant using a dominant test.** Plot points represent the estimated power for a given N (cases+controls) at the effect size (Odds ratio) indicated by color. Figure panels are split column-wise according to the minor allele frequency (MAF) for the variant tested (which can be approximately interpreted as the cumulative frequency at which people have a qualifying variant in a gene-level collapsing analysis) and row-wise by the number of controls sampled per-case. Analysis was performed with the *genpwr* R package (v1.0.4).[21]
